# SARS-CoV-2 Infection Biomarkers Reveal an Extended RSAD2 Dependant Metabolic Pathway

**DOI:** 10.1101/2023.05.08.23289637

**Authors:** Samuele Sala, Philipp Nitschke, Reika Masuda, Nicola Gray, Nathan Lawler, James M. Wood, Joshua N. Buckler, Georgy Berezhnoy, Alejandro Bolaños, Berin A. Boughton, Caterina Lonati, Titus Rössler, Yogesh Singh, Ian D. Wilson, Samantha Lodge, Aude-Claire Morillon, Ruey Leng Loo, Drew Hall, Luke Whiley, Gary B. Evans, Tyler L. Grove, Steven C. Almo, Lawrence D. Harris, Elaine Holmes, Uta Merle, Christoph Trautwein, Jeremy K. Nicholson, Julien Wist

## Abstract

We present compelling evidence for the existence of an extended innate viperin dependent pathway which provides crucial evidence for an adaptive response to viral agents like SARS-CoV-2. We show the in vivo biosynthesis of a family of endogenous cytosine metabolites with potential antiviral activity. Two dimensional Nuclear magnetic resonance (NMR) spectroscopy revealed a characteristic spin-system motif indicating the presence of an extended panel of urinary metabolites during the acute viral replication phase. Mass spectrometry additionally allowed the characterization and quantification of the most abundant serum metabolites showing potential diagnostic value of the compounds for viral infections. In total, we unveiled ten nucleoside (cytosine and uracil based) analogue structures, eight of which were previously unknown in humans. The molecular structures of the nucleoside analogues and their correlation with an array of serum cytokines, including IFN-α2, IFN-γ and IL-10, suggest an association with the viperin enzyme contributing to an endogenous innate immune defence mechanism against viral infection.

## Introduction

Coronavirus disease-2019 (COVID-19) has had profound global impacts on individuals as well as healthcare systems. Thus, the biological properties of the etiological viral agent, severe acute respiratory syndrome-Coronavirus 2 (SARS-CoV-2), have been subjected to unprecedented clinical and scientific scrutiny^1–3^. COVID-19 usually presents with a strong respiratory disease phenotype, combined with variable and complex multifaceted immunological and pathophysiological sequelae^4^ that can impact all major organs, generating a diversity of biochemical and clinical sub-phenotypes^5–7^. The combination of these profound metabolic disruptions coupled with prolonged immune system stimulation leads to persistent manifestations in the form of long-Covid or Post Acute COVID-19 Syndrome (PACS)^8,9^. The systemic complexity of COVID-19 requires multiple integrated analytical platforms to probe effectively the underlying mechanistic processes driving the virus-induced pathological responses. A wide variety of biomarkers have been detected that delineate the onset, progression and recovery of COVID-19 including perturbations in serum/urine amino acids^10–12^, tryptophan metabolites^13–15^, lipids^11,16–18^, lipoproteins^6,19–22^, cytokines^4,23–25^ and other pro-inflammatory markers such as C-reactive protein (CRP), lactate dehydrogenase (LDH) and ferritin^26–29^. Hence, measurement of viral-induced metabolic phenoconversion^30^ provides indices of multiple systemic sequelae that give insight into the pathological processes involved. Metabotyping the COVID-19 patient journey has proved to be key to understanding the immuno-metabolic consequences of SARS-CoV-2 infection. In addition to gauging the clinical and metabolic impact of SARS-CoV-2 infection, it is important to enhance our capabilities for rapid viral testing to address the extent and rate of disease transmission. There is also still a need for discovery of early markers that accurately define the timeframe of viral infections and enhance the ability to detect active viral replication and related patient infectivity.

Humans and other organisms have evolved pathway capacity for the production of endogenous antiviral agents^31,32^. An example of this, is the dephosphorylated form of the nucleoside ddhCTP, 3’-deoxy-3’,4’-didehydro-cytidine (ddhC), which has been detected in human blood serum of COVID-19 patients at an early stage of SARS-CoV-2 infection^33^. From a structural viewpoint ddhC is an analogue of clinically-approved antiviral drugs^34^. Notable small molecule antiviral examples of nucleotide analogues include the DNA polymerase inhibitor acyclovir, administered for the treatment of *Human alphaherpesvirus 1^35^,* as well as the RNA-dependent RNA polymerase inhibitor remdesivir^36,37^ and the RNA-dependent RNA polymerase incorporated mutagen Molnupiravir^38^, both clinically approved for the treatment of SARS-CoV-2 infection^39^. Furthermore, the human (Virus Inhibitory Protein, Endoplasmic Reticulum Associated, IFN-inducible (Viperin) protein, also known as RSAD2 (radical S-adenosyl methionine domain-containing 2), was recently shown to catalyze *in vitro* formation of ddhCTP *via* hydrogen atom abstraction and subsequent formal dehydration of cytidine triphosphate (CTP)^40^, confirming these natural antiviral defenses. ddhCTP acts as an obligate chain terminator, due to the absence of a hydroxyl at position 3’ of the modified furanose ring system. ddhCTP competes with CTP for selective incorporation by various Flaviviral (Zika virus, West Nile virus, Dengue virus, HCV) RNA-dependent RNA polymerases into nascent strands of viral RNA^40^.

Mammalian viperin appears to be key to these viral defenses. Homologues of the viperin enzyme have been discovered across all three domains of cellular life, with fungal, bacterial and archaeal homologues additionally catalyzing the formation of ddhUTP and 3’-deoxy-3’,4’-didehydro guanosine triphosphate (ddhGTP) from UTP and guanosine triphosphate (GTP) respectively^41,42^. Mammalian viperin has also been shown to catalyze the formation of 3’-deoxy-3’,4’-didehydro uridine triphosphate (ddhUTP) from uridine triphosphate (UTP), albeit having an *in vitro* substrate preference of CTP over UTP of approximately one thousand-fold under physiological conditions^43^. This suggests that these viral inhibitory mechanisms are ancient (the conservation of functional viperin homologues across all three domains and numerous bacterial phyla allows a conservative estimate of >3.4 billion years^44,45^) and subsequently have been functionally tuned by evolution. The presence of antiviral nucleosides in human body fluids points to an innate immune capability that is relevant to our understanding of SARS-CoV-2 and other viral infections in man. An understanding of the immuno-metabolic processes related to these nucleosides is essential to allow their assessment as diagnostic markers for viral infections. In order to evaluate ddhCTP as a viral biomarker and to further understand its potential role in the metabolic response to acute SARS-CoV2 infection, we applied a multilevel spectroscopic approach utilizing NMR spectroscopic and mass spectrometric methods to ascertain whether the antiviral ddhCTP is measurable in urine. We detected 10 pyrimidine nucleoside derivatives in human urine of which 8 of them, to our knowledge, are previously unreported. This indicates a more profound metabolic and evolutionary adaptation to viral infection than has previously been suspected.

## Results

### Acute SARS-CoV-2 infection is associated with a distinct urinary metabolic phenotype

Modeling of urine spectral data from 273 SARS-CoV-2 infected individuals (recruited between July 2022 and January 2023 in an ambulatory setting in the metropolitan region of Heidelberg, Germany (Heidelberg Medical University Ethics commission approval: S-324/2020)) and 77 non infected individuals (controls collected at our facility (Murdoch University Human Research Ethics Committee (Approval 2020/053 & 2021/049)) using PCA and OPLS-DA showed a clear metabolic signature of SARS-CoV-2 infection driven by multiple signals in the ‘diagnostic’ chemical shift region ∼δ_H_ (ppm) 6.22-6.36 (Figure 1a-c and S1A-B). This narrow spectra region was taken and an OPLS-DA predictive model^46^ was built with an AUROC of 0.96 (Figure S1C-D). Expansion of this region contrasted for healthy control samples versus the SARS-CoV-2 infected cohort (Figure 1b and 1c respectively) show two distinct spectral motifs (δ_H_ 6.28-6.30, δ_H_ 6.31-6.32) for the infected participants, and a third minor motif at δ_H_ 6.25-6.27. Detailed evaluation of individual 1D spectra revealed four key doublets: δ_H_ 6.29, J = 1.9 Hz; δ_H_ 6.31, J = 2.1 Hz; δ_H_ 6.29, J = 1.9 Hz; δ_H_ 6.26, J = 2.0 Hz stemming from four metabolites (Figure 1d, representing four sets of doublets assigned to metabolites **1**-**4,** detailed information in the supplementary information). Comparison with in-house and public spectral ^1^H-NMR libraries yielded no match for the observed peaks, but their highly specific chemical shifts in combination with their small coupling constants (∼2 Hz) suggested they are derived from an anomeric proton such as that commonly encountered at the CH-1’ position of nucleosides^47^.

**Figure 1:**
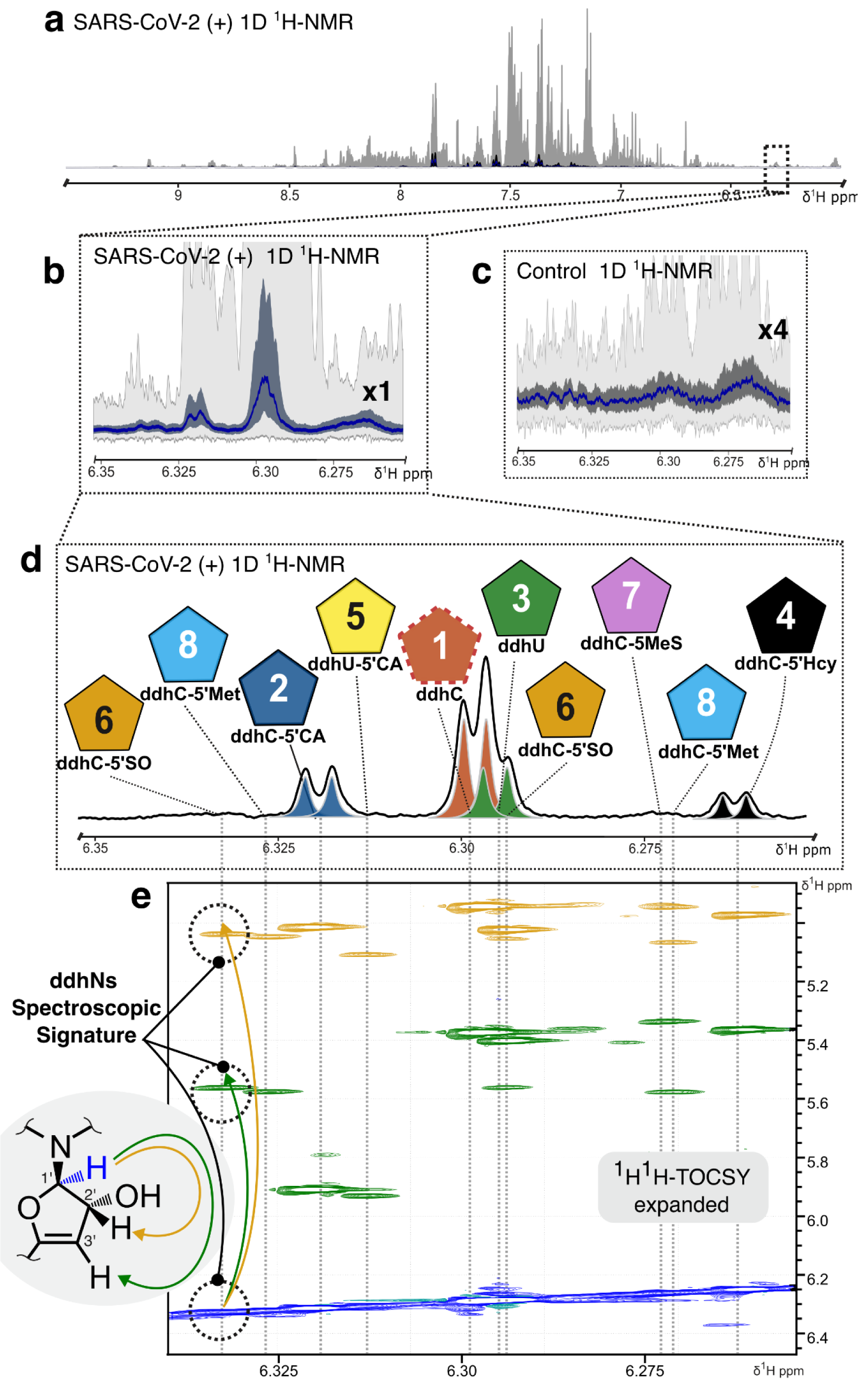
^1^H-NMR metabolic fingerprints of urine from patients with acute SARS-CoV-2 infections: a) Statistical projection of 273 proton NMR spectra from the SARS-CoV-2 (+) cohort focussing on the high frequency part (6-9.5 ppm) of the NMR spectrum. The median NMR spectrum (blue line) is shown with projected upper (Q3) and lower (Q1) quartiles (defined by dark gray area) and the maximum and minimum spectral intensities (light gray). b) Expanded spectral window of the SARS-CoV-2 (+) cohort focussing on the region δ_H_ 6.24-6.35 ppm. Participants from the infected cohort show multiple patterns. c) Comparative spectral window of the projected NMR spectra from the control cohort focussing on the region of interest (ROI). No distinct peaks can be found and the spectral region is dominated by noise. d) Exemplary proton urine NMR spectrum of a SARS-CoV-2 (+) participant depicting the ROI. Four doublets stemming from four different molecular species can be discerned (two of the doublets are overlapping; the individual lines are shown as colored Lorentzian line shapes), which can be assigned to the anomeric protons of the four nucleoside species, 3’-deoxy-3’,4’-didehydro-cytidine (ddhC, **1**), 3’,5’-dideoxy-3’,4’-didehydrocytidine-5’-carboxylic acid (ddhC-5’CA, **2**), 3’-deoxy-3’,4’-didehydro-uridine (ddhU, **3**) and 3’,5’-dideoxy-3’,4’-didehydrocytidine-5’-homocysteine (ddhC-5’Hcy, **4**). The additional metabolites 3’,5’-dideoxy-3’,4’-didehydrouridine-5’-carboxylic acid (ddhU-5’CA, **5**), 3’,5’-dideoxy-3’,4’-didehydrocytidine-5’-methylsulfoxide (ddhC-5’SO, present as a mixture of epimers **6** and **epi-6**), 3’,5’-dideoxy-3’,4’-didehydrocytidine-5’-methylsulfinyl (ddhC-5’MeS, **7**), and 3’,5’-dideoxy-3’,4’-didehydrocytidine-5’-Methionine (ddhC-5’Met, present as a mixture of epimers **8** and **epi-8**) were detected in fractionated urine and dotted lines indicate their anomeric proton chemical shift in urine; **5**, **6**, **epi-6, 7**, **8** and **epi-8** are usually below the limit of detection in standard urine ^1^H-NMR acquisition (∼32 scans, 600 MHz). e) ^1^H^1^H-TOCSY spectrum of the nucleoside “fingerprint” region for the same subject linking the anomeric proton H-1’ (blue) to the full ddhN characteristic allylic H-2’ (yellow) and vinylic H-3’ (green) positions of the dehydrated furanose spin system. In total, 10 ddhN spin systems could be detected.

### Candidate biomarkers of SARS-CoV-2 infection were elucidated

A combination of 800 MHz homonuclear and heteronuclear 2D NMR spectroscopy (Figure S2 and S3), positive-mode HR-MS (QToF), and UHR-MS (FT-ICR) analysis, were applied to urine samples to effect structure elucidation of candidate molecules as follows. The highest intensity peak of metabolite structure of ddhC (**1**) (Figure 1d) could be elucidated and the detected chemical shifts were consistent with published ^1^H- and ^13^C-NMR data (Table S1A and S2). Using a similar approach, the structures of metabolites **2-4**, were elucidated as 3’-deoxy-3’,4’-didehydrocytidine-5’-carboxylic acid (ddhC-5’CA, **2**), 3’-deoxy-3’,4’-didehydrouridine (ddhU, **3**) and 3’,5’-dideoxy-3’,4’-didehydrocytidine-5’-homocysteine (ddhC-5’Hcy, **4**). To our knowledge, metabolites **2** and **3** have not previously been observed. Metabolite **4** (ddhC-5’Hcy) has previously been reported in a mouse urine model of severe *Plasmodium berghei* infection^47^ (Figure 1d). In order to confirm unambigiously the identity of metabolites **1**-**4**, synthetic standards of each were prepared and used in spiking experiments which confirmed the structure and presence of metabolites **1-4** in urine (Figure S5 and S6).

In addition to facilitating structural elucidation of metabolites **1-4**, the high resolution NMR spectra (2D ^1^H^1^H-TOCSY) also indicated the presence of a further six compounds (referred to as deoxy-didehydronucleosides or ddhN with similar but weaker spectroscopic signatures of anomeric H-1’ protons that were not easily detectable in the 1D NMR profiles of urine. Preparative LC fractionation was used to determine the chemical structures of six (**5-8**) additional metabolites (Figure 1d, 1e and S4): 3’-deoxy-3’,4’-didehydrouridine-5’-carboxylic acid (ddhU-5’CA, **5**), 3’,5’-dideoxy-3’,4’-didehydrocytidine-5’-methylsulfoxide, present as a mixture of epimers at the sulfoxide stereocentre (ddhC-5’SO, **6** and **epi-6**), 3’,5’-dideoxy-3’,4’-didehydrocytidine-5’-methylsulfinyl (ddhC-5’MeS, **7**), and 3’,5’-dideoxy-3’,4’-didehydrocytidine-5’-methionine, also present as a mixture of epimers at the sulfonium stereocentre (ddhC-5’Met, **8** and **epi-8**). A comprehensive description of the full structure elucidation of metabolites **1**-**8** is provided in the supporting information of this manuscript and chemical shifts are compiled in Table S1. Synthetic standards of **6**, **epi-6** and **7** were additionally prepared and used in spiking experiments confirming their structure and presence in urine (Figure S5 and S6).

### Extended metabolic network is actively expressed during viral infection

Literature data indicates that CTP undergoes formal dehydration catalyzed by viperin with consumption of one unit of S-adenosylmethionine to generate ddhCTP^40,48^. Considering the close structural homology between the elucidated compounds, a biosynthetic scheme can be deduced linking metabolites **1-8** to the viperin pathway (Figure 2) and one can envisage that nucleotidase catalyzed hydrolysis produces ddhC (**1**) (at urinary concentrations in excess of 20.0 μM/mM creatinine in acute phase patients). Enzymatic oxidation of ddhC (**1**) at position C-5’ (Figure 2a) would yield ddhC-5’CA (**2**); detected at urinary concentrations ca. 10.0 μM/mM creatinine. Hydrolysis of compound **1** via the action of a deaminase enzyme could also generate ddhU (**3**); detected at urinary concentrations ca. 10.0 μM/mM creatinine. It is also possible that UTP is acting as a substrate for *Hs*-viperin *in vivo,* yielding ddhUTP, with subsequent sequential hydrolysis of the triphosphate group producing metabolite **3**. Here we note the approximate 1000-fold substrate preference of CTP over UTP demonstrated by murine viperin *in vitro^43^*, at odds with the elevated concentrations of the downstream product **3** we detect experimentally in human urine from SARS-CoV-2 infected individuals (approx. 2:1, [**1**]:[**3**]). It is possible that both proposed pathways towards metabolite **3** are active under conditions of infection. The downstream substrate promiscuity of the viperin pathway *in vivo* suggests evolutionary adaptation to an expanded panel of viral pathogens, allowing for differential activity of the ddhN pharmacophore against an extended series of viral RNA polymerases. Subsequent sequential oxidation of ddhU (**3**) is likely to produce ddhU-5’CA (**5**); alternatively, the metabolite may arise via hydrolysis of metabolite **2**.

**Figure 2:**
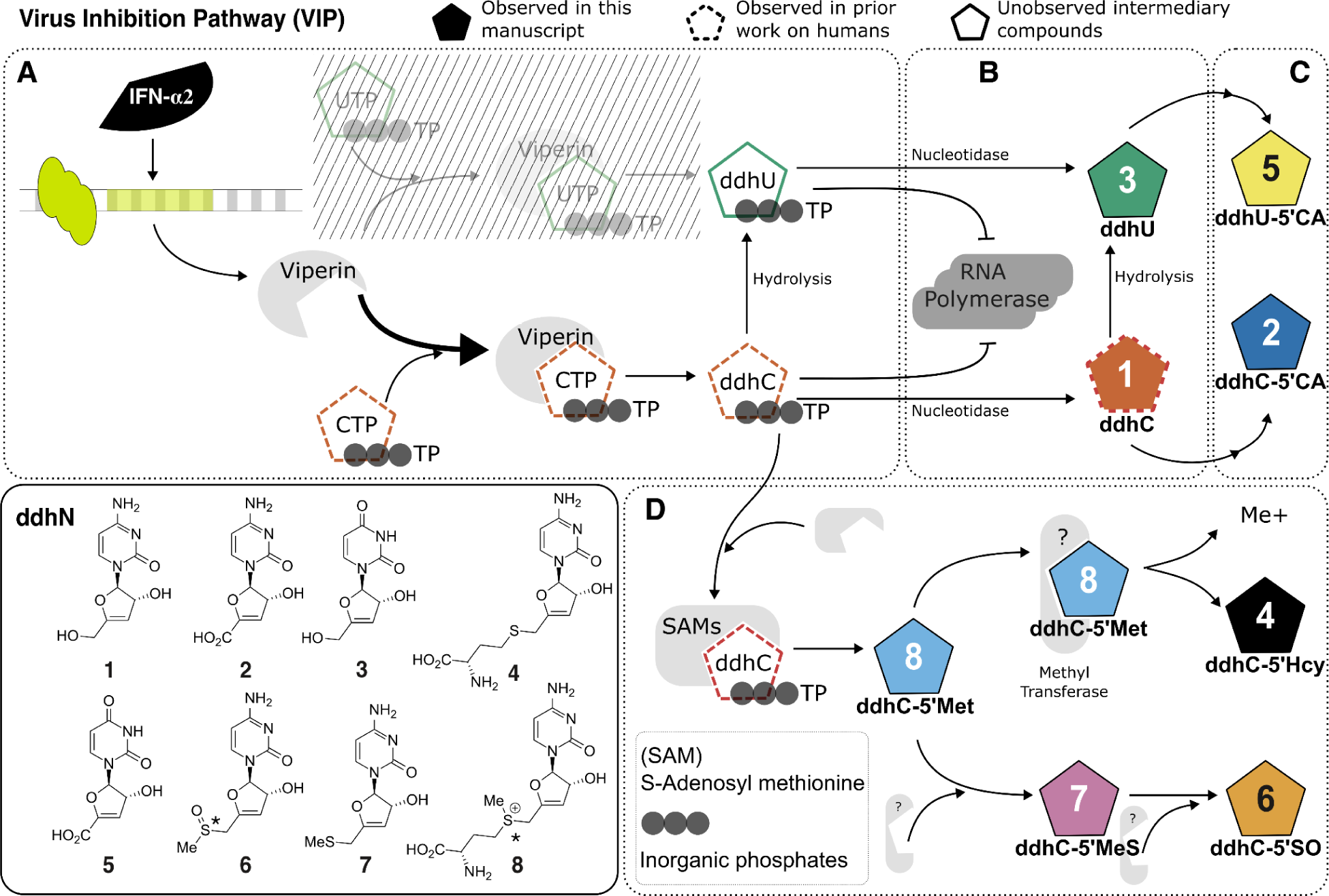
Biosynthetic network leading to the formation of ddhN species. a) Viperin pathway. b) Nucleotidase branch. c) Oxidative Branch. d) Methionine conjugative branch. Figure legend: Color filled pentagons: ddhN metabolites observed in this work; Dashed outlines: metabolites observed or postulated to exist in humans based on prior work; Solid outlines without colour-fill: Unobserved compounds postulated to exist in humans based on this work. Chemical structures of detected ddhN species: ddhC (**1**), ddhC-5’CA (**2**), ddhU (**3**), ddhC-5’Hcy (**4**), ddhU-5’CA (**5**), ddhC-5’SO (**6**), ddhC-5’MeS (**7**) and ddhC-5’Met (**8**).

In the case of the metabolite **4** (detected in urine at concentrations up to 7.0 μM/mM creatinine), a ddhC (**1**) precursor with a suitable leaving group, such as ddhCTP or ddhCDP, could undergo a nucleophilic substitution reaction via an S_N_2-type mechanism, with addition of methionine and concomitant loss of inorganic phosphate to produce ddhC-5’Met, present as a mixture of epimers (**8** and **epi-8**) (Figure 2d). Sulfonium dealkylation via the action of suitable biological nucleophiles would lead to production of urinary metabolite **4**. Parallel work in our laboratories established that S-adenosylmethionine Synthetase (MAT2A isoform) is capable of realizing this reaction sequence (Grove et al. Unpublished results, in redaction). Dealkylation of **8** also yields ddhC-5’MeS (**7**), with subsequent S-oxidation leading to ddhC-5’SO, present as a mixture of epimers (**6** and **epi-6**). The detected relative concentration ratio of metabolite **4**:**7**, approx. 20:1, infers enzymatic, and not chemical, methyl group transfer by native biological methyltransferases^49^, since chemical decomposition would produce byproducts that are not observed. The exact biological functions and implications of this methyl group flux within the biological complex, in a pathway parallel, yet distinct, to SAM methyl group donation, warrants further investigation and is the focus of ongoing investigation in our laboratories.

### Urinary ddhNs derivatives provide diagnostic value in monitoring infection

We investigated the distribution of the ddhN derivatives and assessed their potential as markers for viral infections in urine by NMR comparing samples from SARS-CoV-2 infected patients (n=273) versus non-infected individuals (n=77). Metabolites **4**-**8** were not profiled in detail because they were not detected in all patient samples. Compounds **1**-**3** were directly quantified from the ^1^H-NMR spectra using a peak fitting approach of the anomeric CH-1’ position that was adjusted according to the ERETIC signal^50^, and subsequently normalized to creatinine (Figure 3a and 3b). The results are summarized in Figure 4 and Table S3. By visual inspection of the spectral data, a total of 239 (88%) of SARS-CoV-2 (+) samples were observed to contain ddhC (**1**), of which 55% samples could be reliably quantified. Similarly, ddhU (**3**) were detected in 73% of the SARS-CoV-2 cohort, of which 55% were quantified (Figure 3a), and ddhC-5’CA (**2**) was present in 68% of the samples, of which 34% could be quantified.

**Figure 3:**
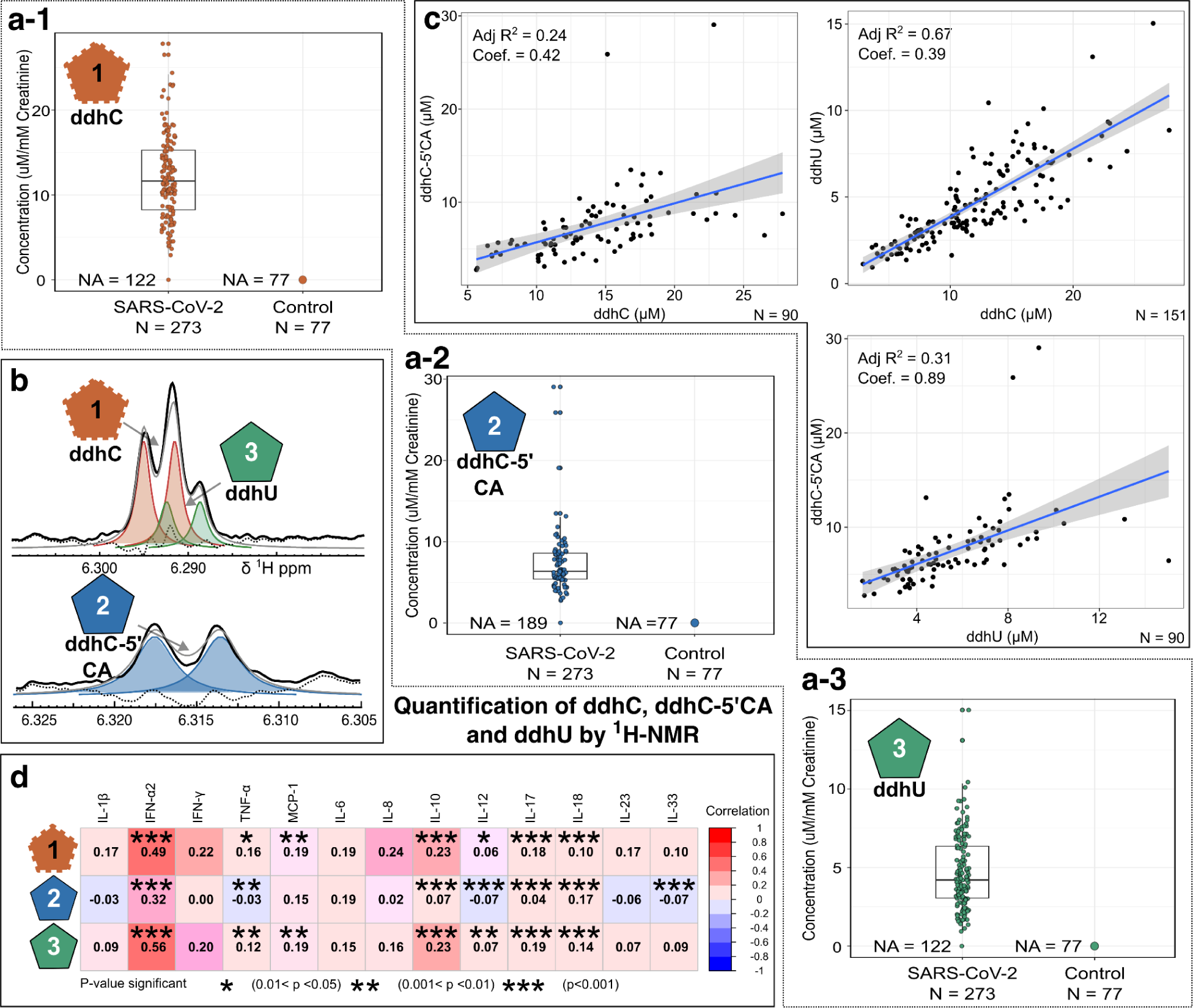
Quantification of ddhN species **1-3** in urine and serum. a) Box plots for compounds **1**-**3** concentration showing the comparison between individuals in the acute phase of SARS-CoV-2 infection and uninfected controls measured by ^1^H-NMR spectroscopy; b) Quantification of 1-3 by spectral deconvolution with colored areas representing the quantities for each metabolites. The black and gray traces represent the experimental and fitted data, while the dashed trace represent the residual; c) Linear regression plots of compound **1**-**3** concentrations measured in urine by NMR. The concentration for all three compounds is given per mM creatinine. All three metabolites were below the limit of detection for all control samples and were not observable following visual inspection of the acquired spectra. d) Pairwise spearman correlation plot between the peak areas for compounds **1**-**3** at the most acute state of SARS-CoV-2 (+) infection in serum measured by LC-QQQ-MS, and cytokines measured via flow cytometry.

**Figure 4:**
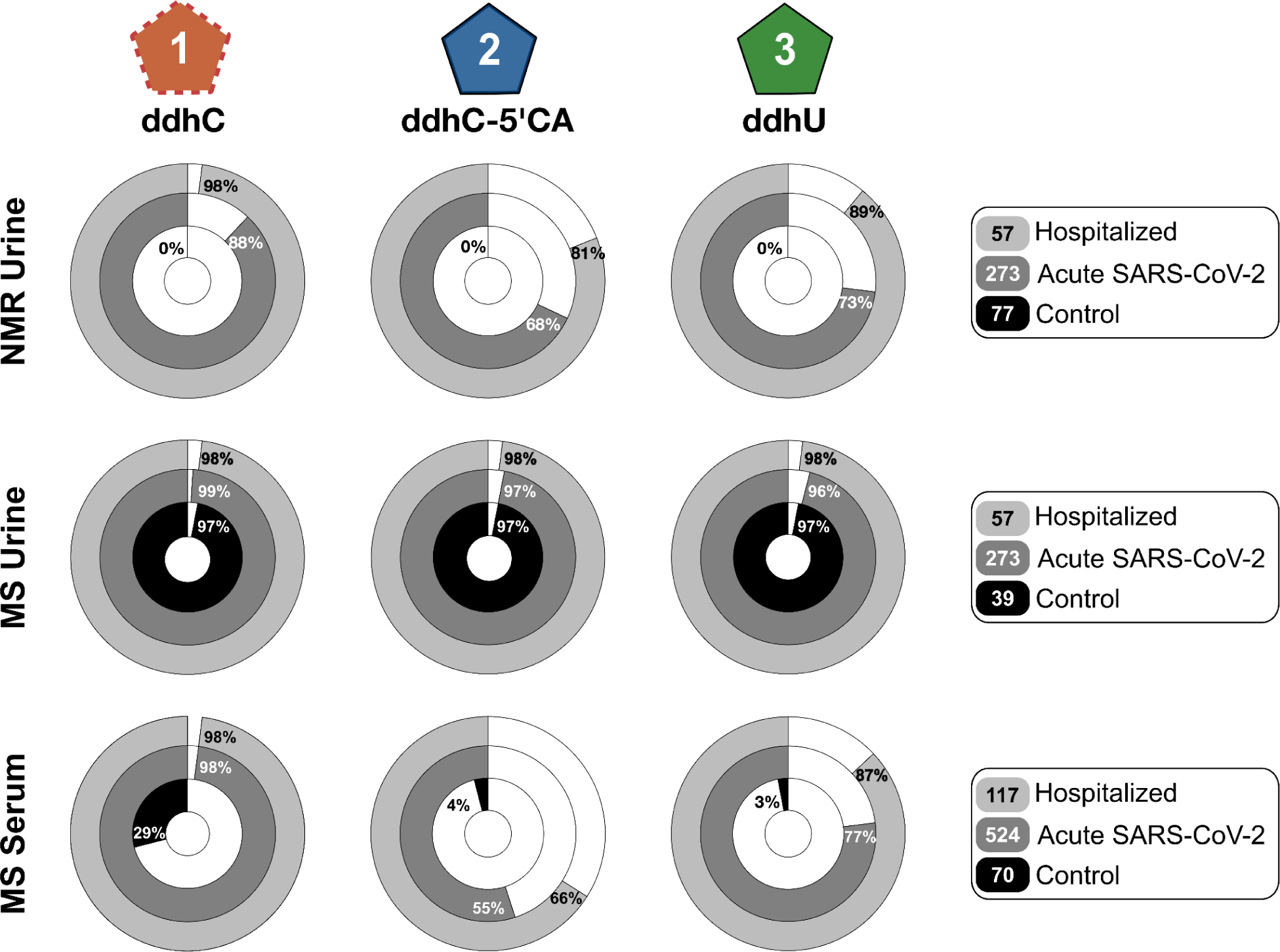
Relative abundance of ddhN species **1-3** in urine and serum as detected by NMR and MS. Left to Right, Top to Bottom, ddhC (**1**) detection by NMR in urine, ddhC-5’CA (**2**) detection by NMR in urine, ddhU (**3**) detection by NMR in urine, ddhC (**1**) detection by QQQ-MS in urine, ddhC-5’CA (**2**) detection by QQQ-MS in urine, ddhU (**3**) detection by QQQ-MS in urine, ddhC (**1**) detection by QQQ-MS in serum, ddhC-5’CA (**2**) detection by QQQ-MS in serum, ddhU (**3**) detection by QQQ-MS in serum. The fact that the three most abundant ddhN derivatives are detected at baseline level in urine but not in serum may be explained by the short half-life time expected for these compounds based on their structural similarity with common antiviral pharmacophores.

In order to evaluate the severity of infection and its impacts on ddhN excretion, the SARS-CoV-2 infected cohort was sub-divided into hospitalized and non-hospitalized (as a proxy for severity). Metabolites **1**-**3** were predominantly detected and quantified in hospitalized patients compared to non-hospitalized participants (Figure 4), whereas ddhN metabolites were not detected in any of the 77 control samples by NMR. Therefore, we propose that measurement of the excretion of ddhN metabolites may be used reliably as direct indication of viral infection and potentially severity^51^.

To probe further how metabolites **1**-**3** are biologically connected, linear regressions were performed (Figure 3c). The correlation (R^2^ = 0.67, r = 0.81) between ddhC (**1**) and ddhU (**3**) indicates that **1** and **3** likely follow the same metabolic step i.e. conversion of CTP and UTP to ddhCTP and ddhUTP by viperin. In contrast, the lower correlations between dhhC (**1**) or ddhU (**3**) with ddhC-5’CA (**2**) (R^2^ = 0.24 and 0.31; r = 0.53 and 0.61 respectively), would suggest that ddhC-5’CA (**2**) might be metabolized downstream in the pathway and/or the conversion rate via oxidative metabolism may be subject to a higher degree of inter-individual variation.

### Time trajectories indicate rapid urinary clearance of ddhNs from the body

For 12% of the participants in the SARS-CoV-2 (+) cohort, none of the nucleoside compounds **1**-**3** could be detected in the urine samples (Table S3). We hypothesized that this may be due to a short half life of the compounds in the body, as some samples were collected up to a week into the acute phase. To test this hypothesis we characterized the excretion profile of nucleosides **1**-**3** over the first seven days post-infection, for six individuals newly infected with SARS-CoV-2 as determined by a positive PCR/RAT test. For all six participants, nucleosides **1**-**3** could be detected from the first sampled time point, indicating a very rapid production of ddhN nucleosides in response to SARS-CoV-2 infection (Figure S8). Typically, the urinary concentrations of compounds **1**-**3** were highest in the first collection post infection with subsequent collections exhibiting a rapid decrease in the concentration of all ddhN metabolites. After 4-5 days of infection, the urinary concentrations of compounds **1**-**3** were close to or below the NMR limit of detection.

### Quantification of ddhN derivatives in urine by LC-QQQ-MS establishes basal viperin expression in healthy controls

In order to quantify urinary ddhNs below their NMR limit of detection, a targeted LC-QQQ-MS analysis was developed in order to quantify nucleosides **1-3** (see supporting information for a complete description of the method). For the LC-QQQ-MS assay, nucleosides **1**-**3** could be detected in 99% of all SARS-CoV-2 infected subjects and 98% of the control cohort (Figure 4). However, the average concentration of all three compounds was at least one order of magnitude higher (13-78 times) in infected participants compared to controls (Figure S9A and Table S5) confirming the potential of ddhNs as urinary biomarkers of SARS-CoV-2 infection and possibly other viral infections.

### Detection of ddhNs in serum is associated with inflammation

The previously reported detection of ddhC (**1**) in the serum of individuals infected with SARS-CoV-2^33^ prompted us to ascertain if ddhC-5’CA (**2**) and ddhU (**3**) are also present in blood. All three nucleosides were detected in serum with the targeted LC-QQQ-MS assay. ddhC (**1**) was present in 98% of all SARS-CoV-2 (+) samples, whereas **2** and **3** were detected in 55% and 77% of samples respectively (Table S5 and Figure S9B). Nucleosides **1**-**3** were also detected in some non-infected control samples (Table S5), but again, in line with the urine results, the average peak area of **1**-**3** in control serum was determined to be one to two orders of magnitude lower compared to infected subjects (Figure S8B and Table S5).

To explore the inflammatory role of viperin-associated ddhNs, serum concentrations of nucleosides **1**-**3** were correlated with cytokine levels. Absolute serum concentrations of 13 cytokines were measured and strong correlations of ddhC (**1**) and ddhU (**3**) were observed with interferon-α2 (IFN-α2). A significant correlation between ddhC-5’CA (**2**) and IFN-α2 was also noted (Figure 3d). This is consistent with previous reports of viperin expression and downstream ddhCTP biosynthesis being induced by type-I interferons^52^.

## Discussion

SARS-CoV-2 infection results in a complex range of immunological and metabolic perturbations across multiple organ systems, which manifest as observable abnormalities in multiple biochemical pathways as reflected in biofluid profiles. In this study, extensive, cohort wide metabolic profiling analysis of urine from acutely infected individuals allowed for the detection of a key spectroscopic signature (Figure 1) attributed to the anomeric CH-1’ proton class of compounds defined as deoxydidehydronucleosides (ddhNs). We observed and elucidated ten discrete ddhN molecular signatures (Seven MSI^53^ level 1 (Figure S5) and three MSI level 2). The most abundant compounds **1**-**3** were successfully quantified in serum samples of the same SARS-CoV-2 infected cohort by means of LC-MS profiling.

Based on the measured concentrations and published literature,^41,42,54^ an elaborate viral inhibitory pathway is proposed, linking compounds **1-8** to the extended viperin pathway and the known CTP and UTP metabolites (Figure 2). A likely route for ddhU synthesis is via nucleotidase hydrolysis of ddhCTP, with subsequent hydrolysis of ddhC (**1**). Alternatively, UTP may be acting as an *in vivo* substrate for viperin, to generate ddhUTP, with subsequent hydrolysis to nucleoside **3**. Oxidation at position C-5’ of the relevant scaffolds is proposed to produce the observed metabolites **2** and **5**, most likely as a means of excretion from the biological system. In addition, the presence of ddhU-5’CA (**5**) suggests a degree of interchangeability between cytosine and uracil motifs across the metabolite series, offering insight into the possible identity of additional metabolites within the ddhN pathway that currently remain below the conventional limit of detection. Of note, we did not detect the known immediate products of the viperin pathway: ddhCTP, ddhCMP and ddhUTP^40,42^, in any of the clinical samples (urine or serum), likely as a consequence of the triphosphorylated metabolites manifesting decreased plasma-membrane permeability thereby limiting their circulation and presence in measured biological fluids. The action of promiscuous nucleotidases present in the cytosol and in plasma could be another contributory factor leading to a short biological half-life of the phosphorylated species. The structural similarity of **1** and **3** to their antiviral counterparts, which have relatively short half-lives when administered to humans, suggests that decline in concentrations with time seen for these endogenous antivirals may reflect the recent, and short term, induction of enzymatic activity resulting from stimulation of the immune system^55,56^. In this respect metabolites **1** and **3** represent potential extracellular reservoirs of the biologically active ddhCTP and ddhUTP pharmacophores, as well as a means of transport via passive diffusion from the site of infection into systemic circulation.

The presence of the homocysteinylated adduct: ddhC-5’Hcy (**4**), accumulated at high concentrations in urine, is at odds with conventional excretory xeno-metabolic pathways, via conjugation of electrophilic cysteine and/or glutathione to reactive sites within the molecule^57^. Methionine conjugation to the ddhCTP scaffold has been observed, yielding the detected intermediate metabolite ddhC-5’Met (**8** and **epi-8**), with subsequent dealkylation to form the major product **4**, as well as the minor metabolite ddhC-5’MeS (**7**). Subsequent oxidation of **7** would result in the production of ddhC-5’SO (**6** and **epi-6**). Conjugation of methionine to the ddhN scaffold and subsequent enzymatic methyl group donation, as observed from the accumulation of the downstream product **4**, versus the presumed intramolecular chemical dealkylation products **6** and **7**, may have profound system-wide implications, as the ‘lost’ methyl group may have epigenetic or metabolic implications.

Serum metabolites **1-3** correlate strongly with cytokine IFN-α2 (Figure 3d) and IFN-γ. Expression of the viperin enzyme, necessary for the production of **1** and the downstream products **2** and **3**, is induced in a number of cell types by type I (α and β), II (γ), and III (λ) IFNs after infection with a number of viruses^52^. Although IFN-γ is a strong inducer of viperin expression in primary macrophages, IFN-α and IFN-β are more effective at inducing viperin expression in a wider range of cell types^52^. IFN-α is elevated in SARS-CoV-2 infection and associated with an increased abundance of several T cell subsets involved in early antiviral responses^58^. Compounds **1** and **3** were also correlated to interleukin-10, and to a lesser extent, with interleukin-17 and interleukin-18. Both IFN-α and interleukin-10 have been shown to be associated with lung injury, predictive of infection severity^59^ and were present at higher concentrations in symptomatic individuals^60^.

Recent reports indicate excision of ddhCTP by the SARS-CoV-2 exoribonuclease nsp14 following incorporation by viral RNA dependent RNA polymerase *in vitro^61^*. In this regard, correlations between patient prognosis and levels of compounds **1**-**8** should be regarded with caution, as metabolites **1** and **3** likely exert no anti-SARS-CoV-2 activity of their own, despite reports of activity against various flaviviral RNA polymerases. Rather, the metabolites act as biological probes, i.e. metabolic surrogates of virally induced inflammatory response. Further work is necessary to understand the relationship between production of the alkyl-donor: ddhC-5’Met (**8**) and infection severity, as well as long-term patient prognosis.

The major urinary ddhN metabolites **1**-**3** were found to accurately distinguish active SARS-CoV-2 infection from healthy controls (Figure 3a). Quantification was performed by ^1^H-NMR peak fitting, or a targeted LC-QQQ-MS assay using the available standards **1**-**3**. The anomeric protons at position H-1’ of all three compounds resonate in an NMR frequency range that is typically void of other peaks (Figure 1b and c). Hence, quantification of the anomeric proton peak from compounds **1**-**3** is simplified as the peak overlap from other compounds is minimal in this spectral region. NMR spectroscopy of urine presents several advantages as a metabolic profiling platform. These include an extended linear dynamic range, intrinsic absolute quantification and robust instrumentation, and the ease of sample collection and preparation obviating the need for phlebotomists and skilled professionals^62–64^. A recent report by Sriskandan and co-workers reported the detection of ddhC (**1**) in the serum and plasma of SARS-CoV-2 infected individuals experiencing acute viral infection following untargeted HILIC-QToF-MS analysis, with the metabolite shown to outperform white cell counts, lymphocyte counts, and CRP as a viral biomarker.^33^ The authors additionally showed that levels of **1** were 36-fold higher in patients with viral infections, including SARS-CoV-2, in comparison to bacterial infections^33^. Our observation by NMR spectroscopy of metabolites **1**-**8** in the urine of SARS-CoV-2 infected patients represents a number of advantages over the former mass spectrometry based assay, as previously described. Nevertheless, dedicated analyses of ddhNs by means of targeted LC-QQQ-MS in plasma or serum may give additional information and can enable straightforward comparison of ddhNs with classical and contemporary established medical markers (e.g. CRP, LDH, Glyc/SPC ratio) when measured in serum.

Although the cohorts analyzed in this study were restricted to patients infected with SARS-CoV-2, it is expected that the newly observed biomarkers may prove valuable in diagnosing other acute viral infections that may result in clinical actionability with respect to acutely ill patients. In this regard one can draw analogy to the currently employed procalcitonin assay^65^, used for the diagnosis of bacterial bronchitis, pneumonia, COPD exacerbation and severe bacterial sepsis^66–68^. The nucleoside disclosed herein represents a promising development in the search for a general diagnostic assay for acute viral infections with real-world clinical utility^69^.

## Human Ethics

The usage of all samples and data was approved by the Ethics Commission of Heidelberg Medical University (S-324/2020) and the Human Research Ethics Committee (HREC) of Murdoch University (Approval 2020/053, and Approval 2021/049). All participants signed a written informed consent according to the declaration of Helsinki.

## Data availability

The MS and NMR raw data generated in this study have been deposited in the Zenodo data repository: https://zenodo.org/record/7906158

## Data Availability

All data produced in the present study are available upon reasonable request to the authors.
Some data are available online at:

https://zenodo.org/record/7906158

## Acknowledgements

We thank The Spinnaker Health Research Foundation, WA, The McCusker Foundation, WA, The Western Australian State Government and the Medical Research Future Fund (EPCD000037 and MRF2014349) for financial support. We thank the Department of Jobs, Tourism, Science and Innovation, Government of a Western Australian Premier’s Fellowship for RLL and the ARC for Laureate Fellowship funding for EH. JW thanks Ministerio de Ciencia, Tecnología e Innovación (Minciencias), Ministerio de Educación Nacional, Ministerio de Industria, Comercio y Turismo e ICETEX (792–2017) 2a Convocatoria Ecosistema Científico - Colombia Científica para la Financiación de Proyectos de I + D + i), World Bank and Vicerrectoría de Investigaciones, Pontificia Universidad Javeriana, Bogotá, Colombia (contract no. FP44842 - 221-2018). We gratefully thank the Werner Siemens Imaging Center under direction of Prof. Bernd Pichler for supporting this project as well as Aditi Kulkarni and Daniele Bucci for excellent technical assistance. JMW, GBE and LDH thank the New Zealand Ministry of Business Innovation & Employment for support (Endeavour Fund program contract UOOX1904 (NZ)). JKN and JW acknowledge Dr. Oscar Millet for pertinent discussions.

