## Supplementary Information for "SARS-CoV-2 Infection Biomarkers Reveal an Extended RSAD2 Dependant Metabolic Pathway"

**Acknowledgements**: Acknowledgements: We thank The Spinnaker Health Research Foundation, WA, The McCusker Foundation, WA, The Western Australian State Government and the Medical Research Future Fund (EPCD000037 and MRF2014349) for financial support. We thank the Department of Jobs, Tourism, Science and Innovation, Government of Western Australian Premier's Fellowship for funding EH. We thank the ARC for Laureate Fellowship funding for EH. JW thanks Ministerio de Ciencia, Tecnología e Innovación (Minciencias), Ministerio de Educación Nacional, Ministerio de Industria, Comercio y Turismo e ICETEX (792–2017) 2a Convocatoria Ecosistema Científico - Colombia Científica para la Financiación de Proyectos de I + D + i), World Bank and Vicerrectoría de Investigaciones, Pontificia Universidad Javeriana, Bogotá, Colombia (contract no. FP44842 - 221-2018). We gratefully thank the Werner Siemens Imaging Center under direction of Prof. Bernd Pichler for supporting this project and Daniele Bucci for excellent technical assistance.

### Table of Contents

**Table of Contents** **3**
**Materials and Methods** **5**
Chemicals and consumables 5
Cohorts 5
Heidelberg 5
Demographic Table 6
Heidelberg cohort acute urine samples 6
Heidelberg cohort acute serum/plasma samples 7
Healthy control cohort urine sample (NMR spectroscopy) and non-infected cohort for urine and serum (LC-QQQ-MS) 8
Sample preparation, data acquisition and processing 9
1H NMR spectroscopy sample preparation 9
1H NMR spectroscopy data acquisition for (IVDr) 600 MHz 9
NMR spectroscopy data acquisition and processing for 800 MHz 9
Liquid Chromatography, Triple Quadrupole (QQQ) Mass Spectrometry 12
LC-MS/MS Serum sample preparation 12
LC-MS/MS experimental conditions 12
LC-MS/MS Urine sample preparation 13
LC-MS/MS Urine experimental conditions 13
Hydrophobic Interaction Liquid Chromatography, Quadrupole Time-of-Flight Mass Spectrometry (HILIC-QToF-MS) analysis 14
Ultra High Resolution Mass Spectrometry 15
Preparative Liquid Chromatography 16
Cytokine flow cytometry assay 17
Data preprocessing and bioinformatics 18
**Results** **19**
Multivariate modelling using PCA and o-PLS 19
Figure S1. PCA and OPLS models, score and loading plots 19
Structure Elucidation 21
Figure S2. 2D 1H1H-TOCSY, 1H1H-NOESY and 1H13C-HMBC of 1. 22
Figure S3. 2D 1H1H-longrangeCOSY 25
Figure S4. 1D spectra of fractions for compounds 5-8. 29
Figure S5. Spiking of synthetic standards for structural confirmation. 31
Spectra assignments 32
Table S1A. Chemical shifts and assignment of new biomarkers 1-3. 32
Table S1B. Chemical shifts and assignment of new biomarkers 4 and 5. 33
Table S1C. Chemical shifts and assignment of new biomarkers 6 and 7. 34

Table S2. Chemical shift of reference compounds 1 to 3. 35
**Syntheses** **36**
Figure S6: Synthesis of ddhN standards 2,3, 6 and 7 for 1H NMR spike in experiments 36
Table S3. Detection of compounds 1-3 in acute phase SARS-CoV-2 by NMR spectroscopy 41
Table S4. Detection of compounds 1-3 in acute phase SARS-CoV-2 by LC-MS 42
Figure S8. Longitudinal concentration data of compounds 1-4 from the acute SARS-CoV-2 (+) Human urine samples measured by NMR spectroscopy. 43
Figure S9. LC-MS peak area comparison for 1-4 determined in A) urine and B) serum. 44
Table S5 LC-QQQ-MS peak area comparison in urine and serum 45
Figure S10. Area correlation for compounds 1-4 in urine determined by LC-QQQ-MS. 46
Figure S12. MS/MS spectrum for ddhC (1). 48
Figure S13. MS/MS spectrum for ddhC-5'CA (2) 48
Figure S14. MS/MS spectrum for ddhU (3) 49
Figure S15. MS/MS spectrum for ddhC-5'Hcy (4) 49
**References** **50**

### Materials and Methods

#### Chemicals and consumables

Phosphate buffer (1.5 M $KH_2PO_4$, 2 mM $NaN_3$, 0.1% sodium trimethylsilyl propionate-[2,2,3,3-$^2H_4$] (TSP) in $H_2O/D_2O$ 4:1, pH 7.4 ±0.1), and all NMR tubes (5 mm outer diameter SampleJet$^{TM}$ NMR tubes and regular 7 inch, 5 mm NMR tubes) with the corresponding sealing caps were purchased from Bruker A.G. Rheinstetten).

Organic solvents were all Optima-LCMS grade and purchased from Thermo Fischer Scientific (Malaga, WA, Australia). LC-MS grade water was generated from a Milli-Q IQ 7000 (Merck Millipore).

#### Cohorts

#### Heidelberg

Serum and urine samples of the COVID-19 cohort originate from an ambulatory treated-patient collective in the metropolitan region of Heidelberg, Germany. Patients suffering from COVID-19 were cared for on an outpatient basis and their health condition was monitored (Lim et al., 2022). Inclusion criteria of the study were: age ≥ 18 years, SARS-CoV-2 infection detected by RT- PCR, and informed consent for study participation. Exclusion criteria were age ≤ 18 years, inability to use the app, and absence of informed consent.

The usage of all samples and data was approved by the Ethics Commission of Heidelberg Medical University (S-324/2020) and all participants signed a written informed consent according to the declaration of Helsinki.

#### Control Cohorts

A total of 39 urine samples from the 27 in-house bank of healthy cohort participants were used as a part of LC-QQQ-MS “healthy” control. A total of 38 urine samples from the 26 in-house bank of healthy cohort participants were used as a part of NMR spectroscopy “healthy” control. The usage of all samples and data was approved by the Human Research Ethics Committee (HREC) of Murdoch University (Approval 2020/053) and all participants signed a written informed consent according to the declaration of Helsinki.

In addition, a second, control cohort from an in-house collected vaccination study was used (N=500+). From the vaccination study, a total of 39 urine samples from 25 individuals and 70 serum samples from 6 individuals were used to create a “healthy/non-infected” collection of samples and exclude potential reactions of the participants to vaccination. The use of all samples and data was approved by the Human Research Ethics Committee (HREC) of Murdoch University (Approval 2021/049) and all participants signed a written informed consent according to the declaration of Helsinki.

The combination of the in-house healthy bank and vaccination cohort resulted in a total of 77 samples from 51 individuals for the NMR spectroscopic urine analyses, and a total of 39 samples from 27 individuals for the LC-QQQ-MS urine analyses.

For the serum non-infected control cohort, 70 samples of 6 individuals were taken from the vaccination cohort at various time points before and after vaccination.

**Demographic Table**

#### Heidelberg cohort acute urine samples

Total of 273 samples from 273 patients at their first collection time point. Majorities of those were non-hospitalized (n = 186), 21% were hospitalized (n = 57) and some did not have a record of their collection time status (n = 30).

| | **All** N = 273 (100%)[1] | **Non-Hospitalized** N = 186 (68%)[1] | **Hospitalized** N = 57 (21%)[1] | **Non-Specified status** N = 30 (11%)[1] |
|---|---|---|---|---|
| Age (years) | 53.4 (14.6) | 52.7 (14.7) | 58.2 (14.1) | 48.8 (12.4) |
| NA | 1 | 0 | 0 | 1 |
| Gender | | | | |
| Female | 141 / 273 (52%) | 107 / 186 (58%) | 17 / 57 (30%) | 17 / 30 (57%) |
| Male | 131 / 273 (48%) | 79 / 186 (42%) | 40 / 57 (70%) | 12 / 30 (40%) |
| NA | 1 / 273 (0.4%) | 0 / 186 (0%) | 0 / 57 (0%) | 1 / 30 (3.3%) |
| BMI | 28.6 (6.2) | 28.2 (6.4) | 28.9 (5.7) | 30.5 (5.8) |
| NA | 29 | 17 | 8 | 4 |

[1] Mean (SD); n / N (%)

**Heidelberg cohort acute serum/plasma samples**

Total of 337 samples from 337 patients at their first collection time point. Majorities of those were non-hospitalized (n = 231), 20% were hospitalized (n = 67) and some did not have a record of their collection time status (n = 39).

| | **ALL** N = 337 (100%)[1] | **Non-Hospitalized** N = 231 (69%)[1] | **Hospitalized** N = 67 (20%)[1] | **Non-Specified status** N = 39 (12%)[1] |
|---|---|---|---|---|
| Age (years) | 54.1 (15.0) | 52.7 (15.2) | 59.4 (13.6) | 53.4 (14.6) |
| NA | 2 | 0 | 0 | 2 |
| Gender | | | | |
| Female | 177 / 337 (53%) | 132 / 231 (57%) | 24 / 67 (36%) | 21 / 39 (54%) |
| Male | 158 / 337 (47%) | 99 / 231 (43%) | 43 / 67 (64%) | 16 / 39 (41%) |
| NA | 2 / 337 (0.6%) | 0 / 231 (0%) | 0 / 67 (0%) | 2 / 39 (5.1%) |
| BMI | 28.6 (6.0) | 28.4 (6.2) | 29.0 (5.5) | 29.4 (5.7) |
| NA | 30 | 17 | 8 | 5 |

[1] Mean (SD); n / N (%)

Maximum collection time difference within the study participants was 371 days for serum and 85 days for urine.

**Healthy control cohort urine sample (NMR spectroscopy) and non-infected cohort for urine and serum (LC-QQQ-MS)**

For NMR spectroscopy, a total of 77 healthy control urine samples from 51 participants were collected. For LC-QQQ-MS, a total of 39 healthy control urine samples were collected from 27 participants and 70 serum non-infected control samples from 6 participants.

| | **Urine NMR** N = 51 (100%)[1] | **Urine LC-QQQ-MS** N = 27 (100%)[1] | **Serum LC-QQQ-MS** N = 6 (100%)[1] |
|---|---|---|---|
| Age (years) | 36.3 (11.2) | 34.2 (11.4) | 34.6 (8.1) |
| NA | 5 | 2 | 1 |
| Gender | | | |
| Female | 25/51 (49%) | 12 / 27 (44%) | 3/6 (50%) |
| Male | 24/51 (47%) | 13 / 27 (48%) | 3/6 (50%) |
| NA | 2/51 (3.9%) | 2 / 27 (7.4%) | |
| BMI | 24.0 (4.5) | — | 21.1 (2.6) |
| NA | 29 | 27 | 1 |

[1] Mean (SD); n / N (%)

### Sample preparation, data acquisition and processing

#### $^1$H NMR spectroscopy sample preparation

Urine samples or urine after fractionation (see Preparative Liquid Chromatography) were thawed at 4 °C for 2 h then centrifuged for 15 minutes at 13000 g at 4 °C. Urine samples were prepared in 5 mm outer diameter SampleJet™ NMR tubes, following the recommended procedures for *in vitro* analytical and diagnostics procedures[1] using 540 µL of Urine mixed with 60 µL phosphate buffer. Additionally, the samples were sonicated for 5 minutes at ambient temperature prior to analysis.

#### $^1$H NMR spectroscopy data acquisition for (IVDr) 600 MHz

NMR spectroscopic analyses were performed on a 600 MHz Bruker Avance III HD spectrometer, equipped with a 5 mm BBI probe and fitted with the Bruker SampleJet™ robot cooling system set to 5 °C. A full quantitative calibration was completed prior to the analysis using a protocol described elsewhere[1]. All IVDr methods were acquired at 300 K.

The standard one-dimensional (1D) experiments with solvent suppression (pp: noesygppr1d) were acquired with 32 scans (+4 dummy scans), 64k data points, relaxation delay of 4.0 s, and a spectral width of 20 ppm resulting in a total experiment time of 4 min 3s (in line with the Bruker *In Vitro* Diagnostics research IVDR methods).

The spike-in experiments were performed with the same acquisition parameters as the standard 1D experiment at 300 and 320K. The experiments were performed on urine samples that showed "high" amounts of **1-4, 6, epi-6** and **7**. The synthetic standards of **1**-**46, epi-6** and **7** were prepared in 200 µL solutions (~1-5 mg/ml). After data acquisition of the respective sample, increasing amounts (1-28 µL < 5 % volume of initial urine NMR sample) of standard **1**-**46, epi-6** and **7** were directly added to the sample. After thorough mixing the sample was measured again.

Time domain data were Fourier transformed and processed in automation using Bruker Topspin™ 3.6.2 and an exponential line broadening of 0.3 Hz was applied to the 1D water suppressed experiment,

#### NMR spectroscopy data acquisition and processing for 800 MHz

The NMR spectra were acquired on a 800 MHz Bruker AvanceNeo spectrometer, equipped with a 5 mm TCI cryo-probe and fitted with a Bruker SampleJet™ robot cooling system set to 5 °C.

The standard one-dimensional (1D) experiments with solvent suppression (pp: noesygppr1d) were acquired with 32-128 scans (+4 dummy scans), 128k data points, relaxation delay of 4.0 s, and a spectral width of 30 ppm resulting in a total experiment time of 4 min 3s (for 32 scans).

Selective 1D $^1$H-TOCSY spectra were acquired with a standard sequence (pp: seldigpzs; with additional pre-saturation) using a *z*-filter adjusted according to the description made elsewhere[2]. Selective Gaussian pulses for refocusing were determined either using the Bruker ShapeTool or the Bruker interactive interface for selective experiments.. **TD**: 64k; **SW**$_{ppm}$: 20; **D1**: 1.5-2.0 s; **DS**: 4; **NS**: 1k-16k; **Spinlock duration**: 120-180 ms.

Selective 1D $^1$H NOESY spectra were acquired with a standard sequence (pp: selnogpzs.2; with additional pre-saturation) using a *z*-filter adjusted according to the description made elsewhere[2]. Selective Gaussian pulses for refocusing were determined either using the Bruker ShapeTool or the Bruker interactive interface for selective experiments. **TD**: 64k; **SW**$_{ppm}$: 20; **D1**: 1.5-2.0 s; **DS**: 4; **NS**: 1k-16k; **Mixing time**: 800 ms.

$^1$H$^1$H-COSY spectra were acquired with a standard sequence (pp: cosygpprqf). **TD** F1: 4k-8k; **TD** F2: 1k-2k; **SW**$_{ppm}$ F1: 13; **SW**$_{ppm}$ F2: 13; **D1**: 2.0 s; **DS**: 16; **NS**: 2. Due to the long evolution times, some long range couplings are present in the COSY spectra.

$^1$H$^1$H-$_{longrange}$COSY spectra were acquired with a standard COSY sequence (pp: cosygpprqf) as a base, which was slightly modified to incorporate 2 delays (D6) before and after the second 90° hard pulse to allow for an additional coupling evolution of 400 ms . **TD** F1: 4k-8k; **TD** F2: 512-1k; **SW**$_{ppm}$ F1: 13; **SW**$_{ppm}$ F2: 13; **D1**: 2.0 s; **D6** = 200 ms **DS**: 16; **NS**: 4-20.

$^1$H$^1$H-TOCSY spectra were acquired with a standard sequence (pp: dipsi2gpphzs; with additional pre-saturation or pp: dipsi2esgpphzs; usually pre-saturation is preferred due to the additional suppression of urea in urine samples, but sometimes the superior suppression of excitation sculpting was necessary for good spectral quality) using a *z*-filter adjusted according to the description made elsewhere[2]. Parameters: **TD** F1: 4k-16k; **TD** F2: 512-2k; **SW**$_{ppm}$ F1: 13; **SW**$_{ppm}$ F2: 13; **Spinlock duration**: 80-180 ms; **D1**: 2.0 s; **DS**: 32; **NS**: 2-16.

$^1$H$^1$H-NOESY spectra were acquired with a standard sequence (pp: noesyesgpphzs) using excitation sculpting for water suppression and a *z*-filter adjusted according to the description made elsewhere[2]. Parameters: **TD** F1: 8k; **TD** F2: 512; **SW**$_{ppm}$ F1: 12; **SW**$_{ppm}$ F2: 12; **Mixing time**: 800 ms; **D1**: 2.0 s; **DS**: 16; **NS**: 32.

$^1$H$^{13}$C-HSQC spectra were acquired with a standard sequence (pp: hsqcedetgpsisp2.3 or hsqcetgpisp2.3) including multiplicity editing and improved sensitivity scheme[3]. Parameters:

**TD** F1: 4k; **TD** F2: 256-512; $\mathbf{SW}_{ppm}$ F1: 13; $\mathbf{SW}_{ppm}$ F2: 185; $^{1}\mathbf{J}_{(13C1H)}$: 125 or 145 Hz; **D1**: 2.0 s; **DS**: 32; **NS**: 8-32.

$^{1}H^{13}C$ HMBC spectra were acquired with a standard three-fold low-pass J-filter sequence (pp: hmbcetgpl3nd; with additional pre-saturation). Parameters: **TD** F1: 4k; **TD** F2: 128-512; $\mathbf{SW}_{ppm}$ F1: 13; $\mathbf{SW}_{ppm}$ F2: 230; $^{1}\mathbf{J}_{(13C1H)min}$: 120 Hz; $^{1}\mathbf{J}_{(13C1H)max}$: 170 Hz; $^{n}\mathbf{J}_{(13C1H)long\ range}$: 5-8 Hz; **D1**: 2.0 s; **DS**: 16; **NS**: 72-256.

Time domain data were Fourier transformed and processed manually using Bruker Topspin™ 4.1.3, Bruker Topspin™ 4.1.4 and Bruker Topspin™ 4.2.0.

### Liquid Chromatography, Triple Quadrupole (QQQ) Mass Spectrometry

#### LC-MS/MS Serum sample preparation

15 µL of sample was protein precipitated with 85 µL acetonitrile. Samples were vortex mixed for 10 minutes at 4°C and centrifuged for 30 minutes at 4°C. 80 µL supernatant was transferred to a 96-well plate and extracts were dried at room temperature before being reconstituted with 45 µL water. A pooled sample was created using the serum from the total cohort for use as a pooled quality control (PQC) sample throughout analysis. Stock solutions of the standards were prepared at 1 mg/mL in water and diluted to solutions at 1 µg/mL for MS optimisation and retention time confirmation.

#### LC-MS/MS experimental conditions

Reversed-phase chromatographic separation was performed using an Elute™ UHPLC system (Bruker Daltonics, Breman, Germany) on a Waters Acquity BEH C18 column 1.7 µm, 50 x 2.1 mm. Mobile phase A comprised water with 0.5 % formic acid (v/v) and mobile phase B was acetonitrile with 0.5 % formic acid (v/v). A sample volume of 10 µL was injected. Gradient elution was performed at 0.4 mL/min with a starting composition of 5% B held for 0.5 min, increasing to 20 % B at 1.0 min, 95% B at 1.2 min and held for 0.5 min before returning to 5 % B after 1.8 min for a further 2.3 min to ensure re-equilibration. The column oven was held at 60°C and autosampler at 6°C. PQCs and long term references (LTR) were analyzed every 12th injection, monitoring for matrix consistency and measurement consistency. The current assay is fit for purpose and undergoing full validation.

Mass spectrometric detection was performed on an EVOQ tandem quadrupole analyser (Bruker) operated in positive electrospray ionisation (ESI) mode. Source parameters were as follows: spray voltage 4000V; cone temperature 350°C; cone gas flow 20; heated probe temperature 350°C; probe gas flow 40; nebuliser gas flow 40. Multiple reaction monitoring (MRM) was performed for the three analytes of interest with parameters detailed below. Data were acquired using HyStar v5.1 and processed using TASQ 2022 (Bruker Daltonics) resulting in peak area values that were used for downstream analyses.

| Compound | RT (min) | Precursor ion (m/z) | Product ion 1 (m/z) | Collision energy (CE) | Product ion 2 (m/z) | Collision energy (CE) |
|---|---|---|---|---|---|---|
| ddhC (**1**) | 0.66 | 226.1 | 112.1 | 15 | 190.0 | 10 |
| ddhC-5’CA (**2**) | 0.60 | 240.1 | 112.0 | 14 | - | - |
| ddhU (**3**) | 0.70 | 227.1 | 148.0 | 12 | 115.1 | 9 |

**LC-MS/MS Urine sample preparation**

10 µL of sample was diluted with 190 µL water (1:19 v:v) and vortex mixed. A pooled sample was generated using the urine from the total cohort for use as a PQC sample throughout analysis.

**LC-MS/MS Urine experimental conditions**

Reversed-phase chromatographic separation was performed using an Elute™ UHPLC system (Bruker) on a Waters Acquity HSS T3 column 1.7 µm, 100 x 2.1 mm. Mobile phase A comprised water with 0.5 % formic acid (v/v) and mobile phase B was acetonitrile with 0.5 % formic acid (v/v). A sample volume of 5 µL was injected. Gradient elution was performed at 0.3 mL/min with a starting composition of 0% B held for 0.2 min, increasing to 35 % B at 2.0 min, 95% B at 2.1 min and held for 0.4 min before returning to 0 % B at 2.6 min for re-equilibration at 3 min. The column oven was held at 40°C and autosampler at 6°C. PQCs and LTRs were analyzed every 12th injection, monitoring for matrix consistency and measurement consistency. The current assay is fit for purpose and undergoing full validation.

Mass spectrometric detection was performed on an EVOQ tandem quadrupole analyser (Bruker) operated in positive electrospray ionization (ESI) mode. Source parameters were as follows: spray voltage 4000V; cone temperature 350°C; cone gas flow 20; heated probe temperature 350°C; probe gas flow 40; nebuliser gas flow 40. MRM was performed for the analysis of ddhC (1), ddhC-5'CA (2), and ddhU (3) using the parameters detailed in table below. Data were acquired using HyStar v5.1 and processed using TASQ 2022 (Bruker Daltonics).

| Compound | RT (min) | Precursor ion (m/z) | Product ion 1 (m/z) | Collision energy (CE) | Product ion 2 (m/z) | Collision energy (CE) |
|---|---|---|---|---|---|---|
| ddhC (**1**) | 1.60 | 226.1 | 112.1 | 15 | 190.0 | 10 |
| ddhC-5'CA (**2**) | 0.98 | 240.1 | 112.0 | 14 | - | - |
| ddhU (**3**) | 2.20 | 227.1 | 148.0 | 12 | 115.1 | 9 |

**Hydrophobic Interaction Liquid Chromatography, Quadrupole Time-of-Flight Mass Spectrometry (HILIC-QToF-MS) analysis**

**Sample Preparation**

Samples were protein crashed with ice cold Acetonitrile containing internal standards at 1ppm using the following ratio (1:3). Each sample was vortex mixed and then centrifuged at 13,000 x g for 10 mins at 4 °C. The supernatant was collected and transferred to a 96 well plate for analysis. Biological quality control (QC) samples were prepared in an identical manner and injected within the analytical run.

**LC and MS Methods.**

Non-targeted HILIC methodologies were performed using a Waters Acquity I-class UPLC system (Waters Corp, Milford, MA), on a Waters ACQUITY UPLC BEH Amide Column (1.7 µm, 2.1 mm × 100 mm, 130 Å) with a sample injection volume of 3 µL. The mobile phase consisted of A (10 mM of Ammonium Formate in $H_2O$ with 50mM formic acid) and B (10 mM ammonium formate in 90:10 MeCN:$H_20$ with 50mM formic acid) with a constant flow rate of 0.4 mL/min. The gradient began with 100% mobile phase B for 2.45 min and was then linearly increased to 70% from 2.45 to 8.15 min and then to 40% from 8.15 to 9.95 mins. After holding mobile phase B at 40% for 1 min, the percentage was returned to 100% from 10.95 and held until 15 mins. The temperature of the column oven was set at 45 °C. High Resolution Mass spectra were collected using Bruker Impact II quadrupole time-of-flight (QToF) mass spectrometer (Bruker Daltonics) operated with full scan (50 – 1200 m/z) and Auto MS/MS. The ion source settings were: capillary voltage = 4.5 kV; end plate offset = 500 V; drying gas flow = 12.0 L/min; nebulizer gas = 5.0 bar; drying temperature = 250 °C. The data acquisition rate was set to 8 Hz. Low and high energy for MS/MS were set at 20 and 50 eV. An internal calibration was performed by injection of 5 mM sodium formate solution in water:isopropanol (50:50 v/v) at the beginning of every run. The untargeted profiling data were acquired in ESI positive ion mode.

### Ultra High Resolution Mass Spectrometry

#### Sample preparation

Samples (urine or plasma) (10 µL) were diluted 1:50 in Methanol/Water (9:1) 0.1% formic acid, cooled at -20°C for 10 minutes then centrifuged at 15,000 g for 10 minutes at 4°C. The supernatant was collected and transferred into a new vial prior to analysis.

#### Fourier Transform Mass Spectrometry

A Bruker SolariX 7T 2XR Magnetic Resonance Mass Spectrometer was operated in quadrupolar detection mode (2ω), magnitude and positive ionisation mode across the mass range *m/z* 107.5-2000 using Compass ftmsControl (ver. 2.3.0). A data size of 8M, 1M or 512k providing an estimated resolving power of 560k, 70k or 35k@400 *m/z*, 100-200 scans were averaged with an accumulation time of between 0.2-1s used. Atmospheric Pressure Ionisation source settings were Capillary Voltage 4500V, Nebulizer 1.0 bar, Dry Gas 4.0 l/min, Dry Temp. 200°C. Source optics, Capillary Exit 200V, Deflector Plate 220V, Funnel 1 150V, Skimmer 1 15V, Funnel RF Amplitude 120 Vpp; Octopole Frequency 5 MHz, RF Amplitude 350 Vpp; Collision Cell Voltage -2.0V, DC Extract Bias 0.2V, RF Freq. 2 MHz, Collision RF Amp. 1400.0 Vpp, Transfer Optics Time of Flight 0.4 ms, Frequency 6, RF Amplitude 350 Vpp were optimised for low molecular weight.

#### Plasma Screening

For automated injection a Bruker PAL RSI liquid handling robot with 3 drawer Peltier Stack and, LCP 1 Robot Injection Arm and Cheminert Valve Drive, coupled to a Bruker Elute Pump and Column Oven was operated using Compass Hystar (ver. 5.1), a sample loop was loaded with 20 µL prior to injection using a direct infusion method over 3.5 minutes. Sample was delivered to the MRMS using 100% MeOH with the following solvent flow program 0-0.15 min 0.1 mL/min, 0.2-1.95 min 0.01 mL/min, 2.0-2.75 min 0.2 mL/min, 2.75-3.5 min 0.1 mL/min. The solvent was passed through a Waters 2.1mm, 0.2 µm filter frit and assembly prior to nebulisation. The instrument settings were modified, with the quadrupole set to *m/z* 238 using an isolation window of 30 Da, data size was set to 512k. Ions observed in plasma using high-throughput screening: ddhC (**1**) calc. $C_9H_{12}N_3O_4$ $[M+H]^+$ *m/z* 226.082232, obs. *m/z* 226.08224 (ΔmDa 0.008, 0.04 ppm), ddhC-5'CA (**2**) calc. $C_9H_{10}N_3O_5$ $[M+H]^+$ *m/z* 240.061497, obs. *m/z* 240.06172 (ΔmDa 0.22, 0.93 ppm).

#### Direct infusion of Urine samples

For direct infusion of urine extract samples were loaded into a Hamilton 250 µL syringe and infused at a rate of 10 µL/min using the same instrument settings as described above for plasma extracts. Instrument settings were modified with the quadrupole set to the expected ion mass (*m/z* 226,227,240, 250) with an isolation window of 5 Da and data size of 8M. Observed in urine using direct infusion: ddhC (**1**) (calc. $C_9H_{12}N_3O_4$ $[M+H]^+$ *m/z* 226.082232, obs. *m/z* 226.08229 (ΔmDa 0.058, 0.25 ppm), ddhC-5'CA (**2**) calc. $C_9H_{10}N_3O_5$ $[M+H]^+$ *m/z* 240.061497, obs. *m/z* 240.06150 (ΔmDa 0.003, 0.01 ppm), ddhU (**3**) calc. $C_9H_{11}N_2O_5$ $[M+H]^+$ *m/z* 227.066248, obs. *m/z* 227.06636 (ΔmDa 0.112, 0.49 ppm)

**Preparative Liquid Chromatography**

Preparative and analytical optimisation scale HPLC were performed using a Waters Preparative HPLC system based on a Waters 2545 binary gradient module, equipped with a Waters 2998 diode array detector (DAD), a Waters Acquity QDa single quadrupole MS detector operating in ESI-(+) mode, and a Waters 2767 sample manager unit employed as a fraction collector. Analytical work was conducted using a Waters OBD $C_{18}$ reversed phase column (100 mm × 4.6 mm, 5 µm) utilising 10 µL injections, and a flow rate of 1.46 mL/min. Preparative scale HPLC was undertaken with a Waters Sunfire OBD $C_{18}$ reversed-phase column (250 mm × 30 mm, 5 µm) with 600 µL injections at a flow rate of 14.0 mL/min. Solvents were of LC-MS grade.

For preparative scale fractionation of urine from SARS-CoV-2 infected individuals, a sample of urine containing elevated levels of compounds of interest (30 mL) was concentrated *in vacuo*, reconstituted in deionised water (3.0 mL) and subsequently centrifuged (13,000 g, 10 min). The sample was decanted, and separated by preparative LC using an isocratic solvent system of 100% $H_2O$ with 0.1% formic acid (v/v), held for 15 minutes, followed by a gradient of 0.0% MeCN: 100% $H_2O$ to 5.0% MeCN: 95% $H_2O$ with 0.1% formic acid (v/v) over 15 minutes, followed by an increased gradient of 5.0% MeCN: 95% $H_2O$ with 0.1% formic acid (v/v) to 50% MeCN: 50% $H_2O$ with 0.1% formic acid (v/v) over 10 minutes. The 50% MeCN: 50% $H_2O$ (with 0.1% formic acid (v/v)) eluent was then held isocratically for 10 minutes and the column was then re-equilibrated to 100% $H_2O$ with 0.1% formic acid (v/v) over 15 minutes. Fractions were collected in 28.0 mL aliquots and concentrated to dryness *in vacuo*. Fractions were subsequently reconstituted in deionised water (1.0 mL) for NMR spectroscopy and MS analyses, affording semi-purified metabolites **8**; $t_R$ = 0-4 minutes, **epi-8**; $t_R$ = 0-4 minutes, **2**; $t_R$ = 4-6 minutes, **1**; $t_R$ = 6-8 minutes, **epi-6**; $t_R$ = 8-10 minutes, **6**; $t_R$ = 10-12 minutes, **4**; $t_R$ = 18-20 minutes, **5**; $t_R$ = 20-22 minutes, **3**; $t_R$ = 22-24 minutes, and **7**; $t_R$ = 26-28 minutes.

**Cytokine flow cytometry assay**

The quantitative analysis of cytokines was performed only for COVID patients who presented a CRP-value > 10 mg/L at some point during the acute infection. Concentrations of CRP were determined in the clinical laboratory of University Hospital Heidelberg by standard ELISA assays (data not shown here). Cytokine and chemokine (IL-1β, IFN-α2, IFN-γ, TNF-α, MCP-1, IL-6, IL-8, IL-10, IL-12p70, IL-17A, IL-18, IL-23, IL-33) concentrations were determined using the commercially available LEGENDplex Human Inflammation Panel 1 (13-plex) based on V-bottom Plate multiplex assay (#740809; BioLegend). For this purpose, 15 µL of serum were diluted with 15 µL of assay buffer and 25 µL of diluted serum was used for further processing in the assay. Calibration curves were prepared with a 4-fold serial dilution method for all cytokines and chemokines with 7 data points to calculate the actual amount of cytokine and chemokine in each sample. The 25 µL samples and standards were added to a 96-well plate followed by the addition of 25 µL assay buffer and 25 µL capture bead. The plate then was incubated at room temperature for 2 hours (on a plate shaker). The plate was washed after the incubation and 25 µL of biotinylated-detection antibodies were added to each well and incubated for 1 hour. After incubation with detection antibodies, 25 µL of a streptavidin-phycoerythrin (SA-PE) antibody were added to each well and incubated in the 96-well plate for an additional 30 minutes. Finally, the 96-well plate was washed twice before 150 µL of washing buffer were added and samples were transferred to 5 mL flow cytometry tubes. Each sample was analyzed with a multi-colour flow cytometry (FACS BD LRSFortessa, Becton Dickinson, New Jersey, USA). Data acquisition files were transferred to the analysis software (Data Analysis Software Suite; LEGENDplex cloud-based software) and final analysis was performed according to the manufacturer’s instructions.

**Data preprocessing and bioinformatics**

1D NMR spectral urine data were preprocessed in the statistical programming language R. ERETIC correction was applied to all spectra to ensure the observed intensities could be quantified[4,5]. The residual water resonance peak (δ 4.6–6.0) was excised along with chemical shift regions where no signals were observed ($\delta < 0.4$ and $\delta > 9.5$). Spectra were baseline-corrected using an asymmetric least-squares method and spectra were normalised using a probabilistic quotient method where the median spectrum is used as the reference.

Statistical evaluation was performed using principal component analysis (PCA) as an unsupervised method. Supervised multivariate analysis was performed using orthogonal projection to latent structures discriminant analysis (OPLS-DA) using the metabom8 package (version 0.2), obtainable at https://github.com/tkimhofer. Scores plots were constructed to show the differences between the controls and SARS-CoV-2 positive patients.
Logistic regression was performed using the lm function in R. Pairwise Spearmanns' correlations were computed between the cytokines and nucleosides in R using the function Cor and visualised using Corrplot.

NMR spectral lineshape fitting was completed in javaScript language using the packages brukerconverter (https://zenodo.org/record/7714532), nmr-processing (https://zenodo.org/record/7641749), github.com/cheminfo/filelist-utils and github.com/nmr-parser. The two overlapping doublets from ddhC and ddhU were modelled using four lines with estimated parameters of $\delta_H$ = 6.29, J = 1.9 Hz, and $\delta_H$ = 6.285, J = 1.9 Hz. ddhC-5'CA was modelled using two lines (doublet) with estimated parameters of $\delta_H$ = 6.3095, J = 2.0 Hz. Similarly, ERETIC and creatinine signal lineshape were modelled to a singlet (single line). Absolute concentrations were obtained by normalising the area of each signal to the area of the ERETIC peak. Concentrations of the ddhNs are then normalised to creatinine for reporting. The optimization yielded an approximate real limit of quantification of ~20 μM for all three compounds under the applied acquisition conditions (note that the concentrations in the text were normalised to creatinine which leads to calculated values lower than 20 μM).

### Results

#### Multivariate modelling using PCA and o-PLS

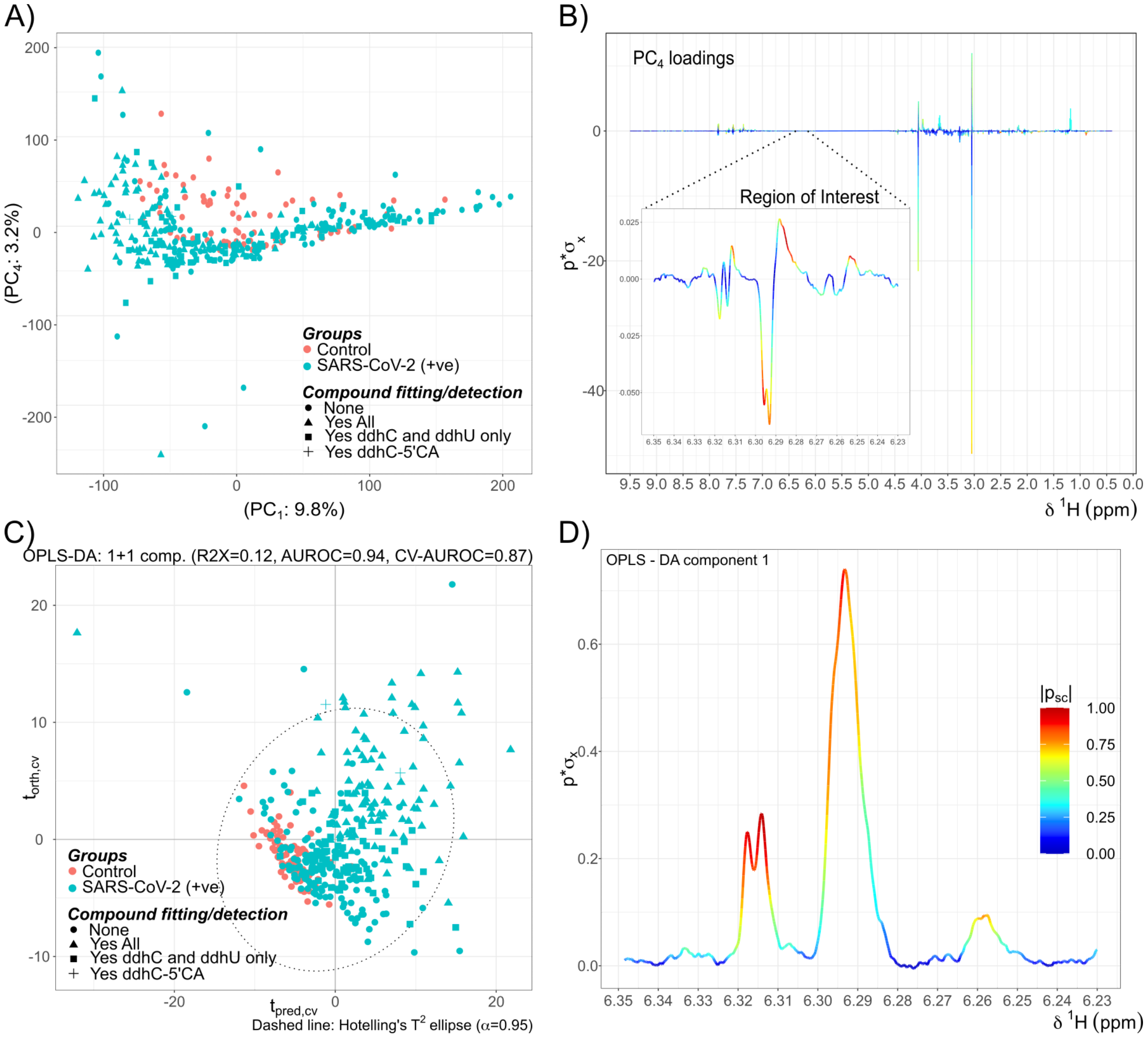


Figure S1. PCA and OPLS models, score and loading plots

Principal components analysis (PCA) and OPLS-DA of urine $^1$H NMR spectra with solvent suppression containing 77control samples and 299 SARS-CoV-2 (+) samples. Baseline correction and probabilistic quotient normalised (PQN) normalisation has been applied prior to the PCA analysis. A) $PC_1$ and $PC_4$ score plot where the compound detection results are represented in different shapes (●: None , ▲: All three compounds ◾ : ddhC anc ddhU only, +: ddhC-5’CA) and their status are indicated by colour (control: orange, SARS-CoV-2 positive: green). B) $PC_4$ loading plot of the whole ppm region (0.5 ~ 9.5 ppm) and the inset shows the loading at the region of interest (6.23 ~ 6.35 ppm). C) OPLS-DA score plot of the $^1$H NMR NOESY spectra at 6.23 ~ 6.35 ppm range. D) corresponding OPLS-DA loading plot.

The loading plot of the fourth principal component (Figure S1B) highlights a strong correlation around 6.293 and 6.296 ppm towards the negative score direction suggesting that those peaks are shown only for the SARS-CoV-2 positive samples. One spectrum from a SARS-CoV-2 (+) subject was excluded due strong spectral deviations and pH induced chemical shifts showing broad peaks throughout the aliphatic range potentially caused by food or drug intake. This resulted in 299/300 spectra for PCA and OPLS analysis.

### Structure Elucidation

### Compound 1

Detailed $^1H$ NMR spectroscopic analysis of metabolite **1** (Urine, 300 K, 800 MHz), aided by combined 2D-$^1H^1H$-COSY and $^1H^1H$-TOCSY NMR spectroscopy permitted assembly of the full spin system: $\delta_H$ 6.02, H-5; $\delta_H$ 7.37, H-6; $\delta_H$ 6.29, H-1'; $\delta_H$ 4.93, H-2'; $\delta_H$ 5.38, H-3'; $\delta_H$ 4.29, H-5'α, $\delta_H$ 4.27, H-5'β (Figure S2A). Salient features of the spin system included a 7.4 Hz coupling between H-5 ($\delta_H$ 6.02) and H-6 ($\delta_H$ 6.37), indicative of an *ortho* $^3J_{H\text{-}H}$ coupling between the protons H-5 and H-6 respectively, and a long range $^5J_{H\text{-}H}$ coupling between proton H-5 and proton H-1' ($\delta_H$ 6.29), discernible via long range $^1H^1H$-COSY analysis and attributed to a particularly favourable W-type geometry between the two protons (Figure S3). $^1H^1H$-NOESY correlations were observed between proton: $\delta_H$ 6.29 (H-1') and protons: $\delta_H$ 4.29, 4.27 (H-5'α/β) as well as proton H-1' and $\delta_H$ 7.37 (H-6) (Figure S2A and C). Multiplicity edited $^1H$-$^{13}C$-HSQC NMR spectroscopic analysis allowed for the assignment of methines: CH-5: $\delta_H$ 6.02, $\delta_c$ 99.4; CH-6: $\delta_H$ 7.37, $\delta_c$ 143.7; CH-1': $\delta_H$ 6.29, $\delta_c$ 96.5; CH-2': $\delta_H$ 4.93, $\delta_c$ 81.4; CH-3': $\delta_H$ 5.38, $\delta_c$ 103.5, and the methylene group $CH_2$-5': $\delta_H$ 4.29, 4.27, $\delta_c$ 58.6. Additional carbon resonances discernible via 2D-$^1H^{13}C$-HMBC NMR spectroscopic analysis (Figure 2D and 2E) included correlations from $\delta_H$ 6.02 (H-5) to the carbonyl: $\delta_c$ 160.1 (C-2); and amidine: $\delta_c$ 169.1 (C-4) and correlations from $\delta_H$ 6.29, (H-1'), $\delta_H$ 4.93 (H-2'), $\delta_H$ 5.38 (H-3') and $\delta_H$ 4.29, 4.27 (H-5'α/β) to a putative enol-ether: $\delta_c$ 164.1 (C-4'). Consideration of the spectroscopic data presented thus far allowed for the assembly of two partial substructures, namely the cytosine residue A (Figure S2A), and a presumed furanose derived residue B (Figure S2D). Observation of two $^1H$-$^{13}C$-HMBC correlations from the anomeric proton $\delta_H$ 6.29 (H-1') to $\delta_c$ 160.1 (C-2) and $\delta_c$ 143.7 (C-6) allowed for assembly of partial structure AB via a CH-1'-N-1 bond as depicted (Figure S2E). To determine the molecular formula of metabolite **1**, LC-ESI-MS analysis of a sample (that contained all three major metabolites in high abundance according to NMR spectroscopy) was undertaken, monitoring all precursor ions leading to the fragmentation of cytosine ($m/z$ = 112.0511, calculated for $C_4H_6N_3O$), a plausible fragmentation for the presumed substructure AB under positive electrospray conditions, via a cationic pinacol rearrangement-type mechanism.[6] Positive-mode HR-MS analysis of the most abundant metabolite compatible with the spectroscopic and spectrometric data presented, indicated a protonated molecular formula: $[M+H]^+$ of $C_9H_{12}N_3O_4$, ($m/z_{calc}$ = 226.0827, $m/z_{found}$ = 226.0826), this inference was corroborated by analysis of the deprotonated molecular ion under negative electrospray conditions: $[M-H]^-$ of $C_9H_{10}N_3O_4$, ($m/z_{calc}$ = 224.0671, $m/z_{found}$ = 224.0674), allowing for the putative molecular formula of metabolite **1** to be determined as $C_9H_{11}N_3O_4$. Additional validation for the purported molecular formula was provided by ultra high resolution MS measurements via Fourier Transform Ion Cyclotron Resonance Mass Spectrometry (FT-ICR-MS) *vide supra* affording a molecular ion consistent with the calculated molecular mass to 0.04 ppm accuracy (calc. $C_9H_{12}N_3O_4$ $[M+H]^+$ $m/z$ 226.082232, obs. $m/z$ 226.08224 (ΔmDa 0.008, 0.04 ppm). A database search of the ascertained molecular formula allowed for the assignment of **1** as the known metabolite: 3'-deoxy-3',4'-didehydrocytidine (ddhC), for which the reported $^{1}$H and $^{13}$C NMR data were in good agreement with that obtained experimentally[7,8].

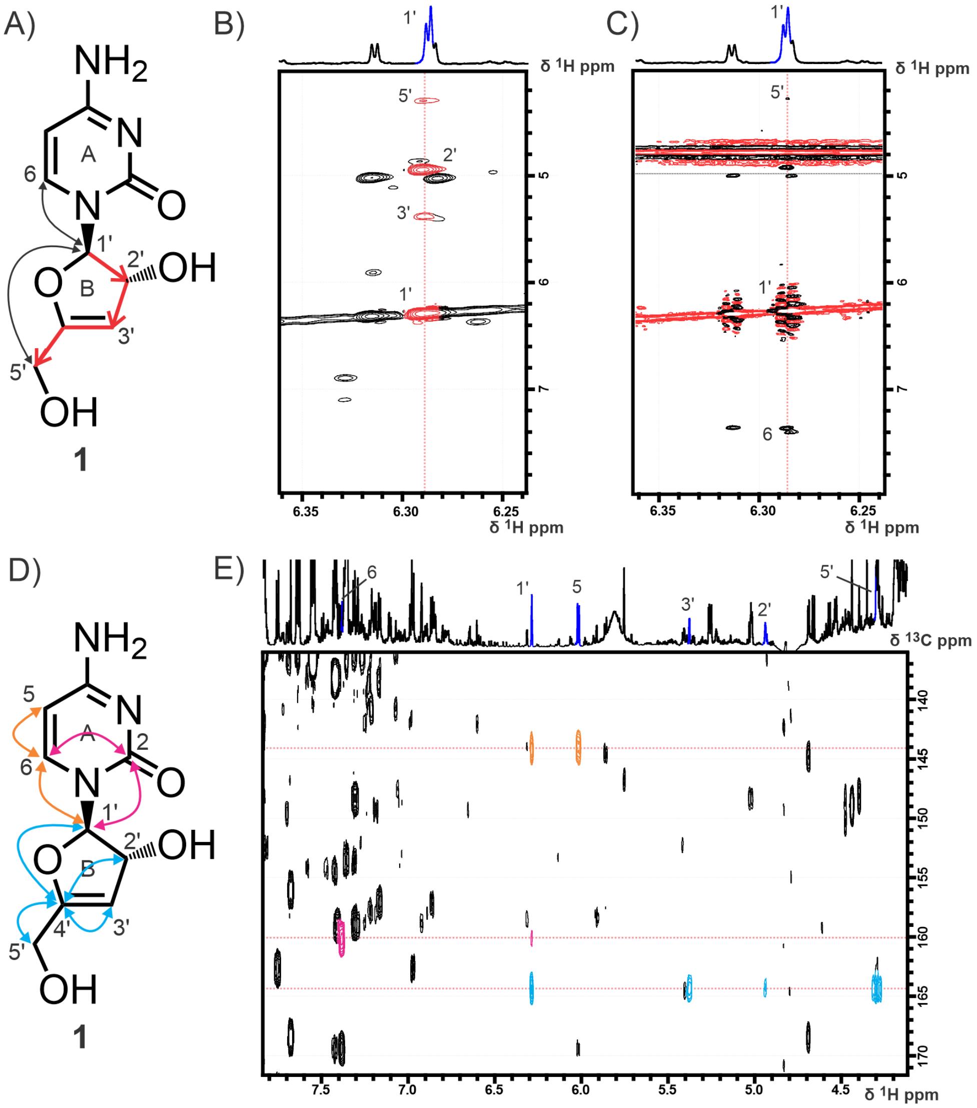


Figure S2. 2D $^{1}$H$^{1}$H-TOCSY, $^{1}$H$^{1}$H-NOESY and $^{1}$H$^{13}$C-HMBC of **1**.
Structural determination of 3'-deoxy-3',4'-didehydro-cytidine (ddhC, **1**) by NMR spectroscopy in urine of a SARS-CoV-2 infected subject during the acute phase. A) Key TOCSY (red arrows) and NOESY (black arrows) correlations, assigning the furanose ring system B and connecting residue A and B. B) Excerpt from a $^{1}$H$^{1}$H-TOCSY spectrum highlighting the spin system originating from H-1' (red) C) Excerpt from a $^{1}$H$^{1}$H-NOESY spectrum highlighting key NOEs from H-1' to H-6 and H-5'. D) HMBC correlations connecting residue A and B (pink and orange

arrows) and key HMBC correlations to position C-4’ (sky blue arrows). E) Excerpt from a $^{1}H^{13}C$ HMBC spectrum highlighting the key HMBC peaks from D.

**Compound 2**

As above, $^{1}H$ NMR spectroscopic analysis of metabolite **2** aided by combined $^{1}H^{1}H$-COSY and $^{1}H^{1}H$-TOCSY NMR spectroscopic analysis allowed for the assembly of the spin system: $\delta_H$ 6.02, H-5; $\delta_H$ 7.37, H-6; $\delta_H$ 6.31, H-1'; $\delta_H$ 5.01, H-2'; $\delta_H$ 5.89, H-3'. Here we note significant spectral overlap between the cytosine fragment of **1** and the presumed cytosine fragment of **2**. Multiplicity edited $^{1}H^{13}C$-HSQC analysis permitted the assignment of the furanose methines CH-1': $\delta_H$ 6.31, $\delta_c$ 97.1; CH-2': $\delta_H$ 5.01, $\delta_c$ 82.4; CH-3': $\delta_H$ 5.89, $\delta_c$ 110.5. $^{1}H^{13}C$-HMBC allowed for the attachment of the cytosine fragment of **2** to the anomeric position CH-1' ($\delta_H$ 6.31), following on from a correlation to position CH-6 ($\delta_H$ 7.37, $\delta_c$ 144.7), as well as allowing for the observation of carbon C-4': $\delta_c$158.8, exhibiting correlations to $\delta_H$ 6.31 (H-1'), $\delta_H$ 5.01 (H-2'), and $\delta_H$ 5.89 (H-3'). Comparison to the spectroscopic data of **1** permitted the discernment of two major points of difference across the respective scaffolds: the disappearance of methylene $CH_2$-5' in compound **2**, as well as an apparent de-shielding of methine CH-3' ($\delta_H$ 5.89, $\delta_c$ 110.5) and shielding of the fully substituted carbon C-4' ($\delta_c$158.8), allowed us to infer the substitution of a hydroxy-methylene at position C-5' of **1** for an electron withdrawing group at the respective position C-4' of compound **2**. LC-ESI-MS analysis of sample 09213-391-1 in positive mode, monitoring precursor ions for a presumed loss of cytidine, and compatibility with the spectrometric data acquired, afforded a protonated molecular formula: $[M+H]^+$ of $C_9H_{10}N_3O_5$, ($m/z_{calc}$ = 240.0620, $m/z_{found}$ = 240.0624). This was further supported by LC-ESI-MS analysis in negative ion mode which afforded a deprotonated molecular ion: $[M-H]^-$ of $C_9H_8N_3O_5$, ($m/z_{calc}$ = 238.0463, $m/z_{found}$ = 238.0469) allowing us to ascertain the molecular formula of **2** as $C_9H_9N_3O_5$. Additional validation for the purported molecular formula was provided via FT-ICR-MS affording a molecular ion consistent with the calculated molecular mass to 0.01 ppm accuracy ($C_9H_{10}N_3O_5$ $[M+H]^+$ *m/z* 240.061497, obs. *m/z* 240.06150 (ΔmDa 0.003, 0.01 ppm). Consideration of the spectroscopic and spectrometric data permitted the assignment of metabolite **2** as 3',5'-dideoxy-3',4'-didehydrocytidine-5'-carboxylic acid, a presumed C-5' oxidized derivative of **1**.

**Compound 3**

Spectroscopic analysis of metabolite **3** was complicated in part by significant overlap of the presumed furanose residue resonances with those of metabolite **1**, nonetheless, a diligent analysis of the combined $^{1}H^{1}H$-COSY and $^{1}H^{1}H$-TOCSY spectra permitted the assignment of the spin system: $\delta_H$ 6.285, H-1'; $\delta_H$ 5.02, H-2'; $\delta_H$ 5.40, H-3'; $\delta_H$ 4.29, H-5'α, $\delta_H$ 4.27, H-5'β. Multiplicity edited $^{1}H^{13}C$-HSQC analysis allowed for the assignment of the furanose methines CH-1': $\delta_H$ 6.285, $\delta_c$ 95.7; CH-2': $\delta_H$ 5.04, $\delta_c$ 81.2; CH-3': $\delta_H$ 5.40, $\delta_c$ 103.2, and the methylene group $CH_2$-5': $\delta_H$ 4.29, 4.27, $\delta_c$ 58.6, suggesting a close analogue of **1**, with postulated modification at the nucleobase moiety of compound **3**. $^{1}H^{13}C$-HMBC analysis indicated a correlation from $\delta_H$ 6.285 (H-1') to $\delta_c$ 144,4 (C-6) as well as a key correlation to $\delta_c$ 154.6 (C-2). Further analysis of the acquired $^{1}H^{13}C$-HSQC and $^{1}H^{13}C$-HMBC spectra permitted the observation of additional methines: CH-5: $\delta_H$ 5.84, $\delta_c$ 105.4; CH-6: $\delta_H$ 7.43, $\delta_c$ 144.4, and the fully substituted carbonyl $\delta_c$ 169.1 (C-4). Consideration of the data at hand, and a comparison to literature chemical shifts indicated the presence of a uracil substructure on compound **3**, attached to the modified furanose residue via C-1'-N-1 as depicted (Fig. 2, main text). LC-ESI-MS analysis of sample 09213-391-1 in positive mode, monitoring for a presumed loss of uracil (*m/z* = 113.0351, calculated for $C_4H_5N_2O_2$), indicated a single feature compatible with the spectroscopic data presented, affording a protonated molecular formula: $[M+H]^+$ of $C_9H_{11}N_2O_5$, ($m/z_{calc}$ = 227.0667, $m/z_{found}$ = 227.0667). HR-MS analysis of the same molecular feature in negative ion mode afforded a deprotonated molecular ion: $[M-H]^-$ of $C_9H_9N_2O_5$, ($m/z_{calc}$ = 225.0511, $m/z_{found}$ = 225.0518), allowing us to conclude that the molecular formula of metabolite **3** was as $C_9H_{10}N_2O_5$. Additional validation for the purported molecular formula was provided via FT-ICR-MS affording a molecular ion consistent with the calculated molecular mass to 0.49 ppm accuracy (calc. $C_9H_{11}N_2O_5$ $[M+H]^+$ *m/z* 227.066248, obs. *m/z* 227.06636 (ΔmDa 0.112, 0.49 ppm). Consideration of the spectroscopic and spectrometric data acquired allowed for the assignment of **3** as the previously reported synthetic: 3'-deoxy-3',4'-didehydrouridine (ddhU). Comparison to the reported $^{1}H$ and $^{13}C$ NMR data of ddhU showed excellent agreement with that of **3**[8].

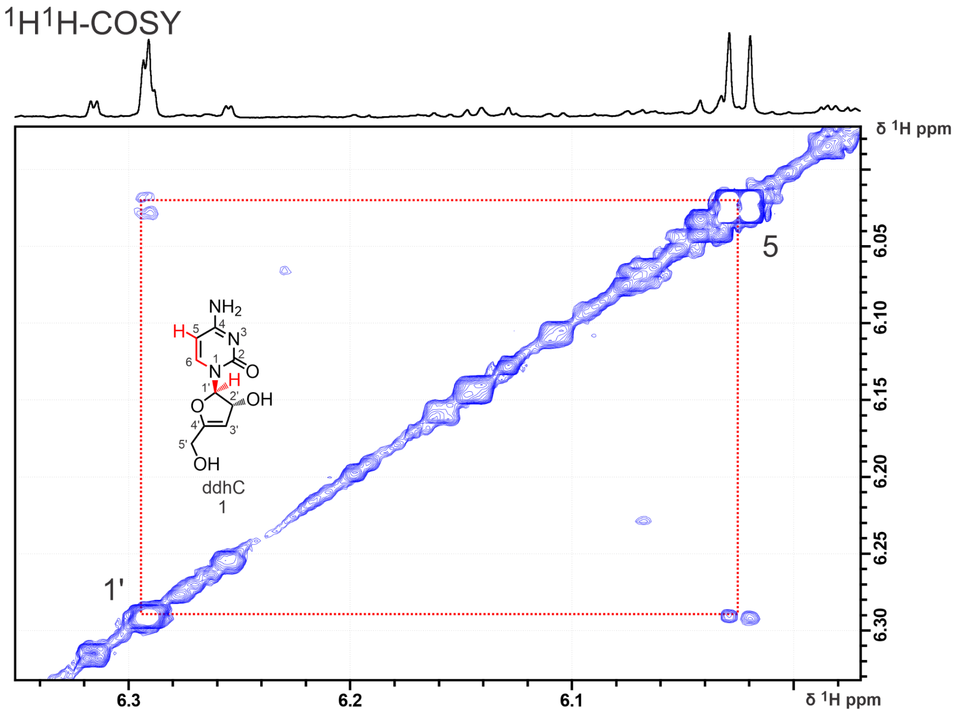


Figure S3. 2D $^1H^1H$-$_{longrange}$COSY

Structural elucidation of ddhC (**1**). $^1H^1H$-COSY highlighting the frequency region $\delta_H$ 5.98-6.35. Clear cross peaks can be seen for the peaks at δ 6.28 and 6.02 representing the long range coupling between the 1' and 5 position of ddhC. Although the acquisition conditions did not directly favour long range couplings, they can still sufficiently evolve due to the high number of indirect points (1024) leading to long $t_1$ evolution delays.

**Compound 4**

Spectroscopic analysis of compound **4** revealed a number of spectral features in common with metabolites **1-3.** An analysis of the combined $^1H^1H$-COSY, $^1H^1H$-TOCSY and $^1H^{13}C$-HSQC spectra obtained from unfractionated acute phase urine indicated a correlation from the anomeric proton H-1' ($\delta_H$ 6.25) to the presumed cytosine residue common to the co-metabolites **1** and **2**: $\delta_H$ 7.46, $\delta_C$ 144.1, CH-6; $\delta_H$ 6.04, $\delta_C$ 99.4, CH-5, and to a dehydrated furanose system: $\delta_H$ 4.97, $\delta_C$ 81.7, CH-2'; $\delta_H$ 5.37, $\delta_C$ 104.6, CH-3'; $\delta_H$ 3.50, $\delta_H$ 3.46, $\delta_C$ 29.7, $CH_2$-5'. The observation of a dehydrated furanose was corroborated by the observation of a $^1H^{13}C$-HMBC cross peak from H-3' ($\delta_H$ 5.37) to position C-4' ($\delta_C$ 161.0). Of note, from this cursory analysis, was the atypically low chemical shift of both the protons and carbon of methylene $CH_2$-5': $\delta_H$ 3.50, $\delta_H$ 3.46, $\delta_C$ 29.7, indicative of a heteroatomic substitution at this position by an element other than oxygen. Complete elucidation of compound **4** was complicated by the extreme spectral overlap of methylene $CH_2$-5' with confounding isochronous carbohydrate resonances in this region of the spectrum, precluding the observation of additional resonances attributable to compound **4** beyond this position and constituting an effective “spin-barrier” for the molecule. To this end, fractionation of a concentrated sample of acute phase SARS-CoV-2 infected urine, separated via preparative HPLC (*vide supra*), afforded a semipurified fraction containing enriched compound **4**, *sans* carbohydrates, for additional analysis. Here, $^{1}H^{1}H$-NOESY analysis revealed an intramolecular NOE from $CH_2$-5' to $\delta_H$ 2.72. Subsequent selective 1D $^{1}H$-NOESY analysis originating from $\delta_H$ 2.72 indicated the presence of a whole spin system H-1'': $\delta_H$ 3.85, $H_2$-2'' $\delta_H$ 2.13-2.20, $H_2$-3'' $\delta_H$ 2.72, suggestive of a homocysteine conjugation to position $CH_2$-5' of **4**. This was corroborated by additional $^{1}H^{1}H$-TOCSY, $^{1}H^{13}C$-HSQC and $^{1}H^{13}C$-HMBC analyses that indicated the full partial substructure: CH-1'':$\delta_H$ 3.84, $\delta_C$ 56.8, $CH_2$-2'' $\delta_H$ 2.19, $\delta_H$ 2.15, $\delta_C$ 32.9, $CH_2$-3'' $\delta_H$ 2.72, $\delta_C$ 29.5. LC-ESI-MS analysis of the sample containing enriched **4** in positive mode, again monitoring precursor ions for a presumed loss of cytidine, and compatibility with the NMR spectroscopic data acquired, afforded a protonated molecular formula: $[M+H]^+$ of $C_{13}H_{19}N_4O_5S$, ($m/z_{calc}$ = 343.1070, $m/z_{found}$ = 343.1071). Consideration of the spectroscopic and spectrometric data allowed for the assignment of **4** as 3',5'-dideoxy-3',4'-didehydrocytidine-5'-homocysteine (ddhC-5'Hcy). The absolute configuration at position CH-1'' of the homocysteine residue is here inferred as S, on biosynthetic grounds.

**Compound 5**

Compound **5** could not be directly assigned from urine due to low signal to noise and spectral overlap. Therefore, a concentrated sample of acute phase SARS-CoV-2 infected urine was fractionated via preparative HPLC. The fractionation yielded a clean fraction that showed almost no additional peaks in the regions of interest for compound **5**. Simple evaluation of the 1D proton spectrum confirmed by $^{1}H^{1}H$-TOCSY yielded the reminiscent dehydrated furanose spin system methines CH-1': $\delta_H$ 6.31, CH-2': $\delta_H$ 5.11 and CH-3': $\delta_H$ 5.93 similar to ddhC-5’CA (**2**). A 1D NOESY spectrum selectively refocusing the methine resonance at $\delta_H$ 6.31 ppm yielded through space connectivity to a doublet at $\delta_H$ 7.43, which in turn could be connected to another doublet at $\delta_H$ 5.87 by $^{1}H^{1}H$-COSY matching the CH-6 and CH-5 positions of a uracil moiety. LC-ESI-MS analysis of the sample containing enriched **5** in positive ion mode, monitoring precursor ions for a presumed loss of uracil, and compatibility with the NMR spectroscopic data acquired, afforded a protonated molecular formula: $[M+H]^+$ of $C_9H_8N_2O_6$, ($m/z_{calc}$ = 241.0453, $m/z_{found}$ = 241.0455). Given the high similarity of the chemical shifts compared to ddhC-5’CA for the dehydrated furanose system, the presence of uracil residue, the apparent absence of a 5' proton peak and high-resolution mass spectral data led to the putative assignment of **5** as 3',5'-dideoxy-3',4'-didehydrouridine-5'-carboxylic acid (ddhU-5'CA, **5**)

**Compound 6**

As with compound **5**, compound **6** also required initial fractionation of acute phase urine in order to increase metabolite concentration, and to reduce the number of confounding resonances complicating structural elucidation of the compound. Analysis of the $^1H^1H$-TOCSY spectrum allowed for the construction of the dehydrated furanose system reminiscent of the homocysteine conjugate (ddhC-5'Hcy, **4**), with key resonances at $\delta_H$ 6.33, CH-1', $\delta_H$ 5.05, CH-2'; $\delta_H$ 5.56, CH-3'; $\delta_H$ 4.00, $\delta_H$ 3.87, $CH_2$-5'. Connection to the cytosine spin system was enabled by simple integration and $^1H^1H$-COSY due to a highly purified fraction in the chemical shift region of interest yielding $\delta_H$ 7.47, CH-6; $\delta_H$ 6.04, CH-5.
Selective 1D $^1H$-NOESY irradiating the methylene $CH_2$-5' in turn revealed a through space correlation to the methyl group: $\delta_H$ 2.81, $CH_3$-1'', which was complemented by $^1H^{13}C$-HSQC: $CH_3$-1'' ($\delta_H$ 2.81, $\delta_C$ 39.6). Finally, LC-ESI-MS analysis of the sample containing enriched **6** in positive mode, monitoring precursor ions for a presumed loss of cytidine, and compatibility with the NMR spectroscopic data acquired, afforded a protonated molecular formula: $[M+H]^+$ of $C_{10}H_{14}N_3O_4S$, ($m/z_{calc}$ = 272.0702, $m/z_{found}$ = 272.0700). Consideration of the spectroscopic and spectrometric data allowed for the putative assignment of **6** as 3',5'-dideoxy-3',4'-didehydrocytidine-5'-methylsulfoxide (ddhC-5'SO, **6**). We note that the relative configuration of the sulfoxide stereogenic centre remains to be determined. There are indications of the epimer of **6** in the fractionated urine with an observed spin system: $\delta_H$ 6.29, CH-1'; $\delta_H$ 5.07, CH-2'; $\delta_H$ 5.56, CH-3', $\delta_H$ 3.77, $CH_2$-5' deemed **epi-6**, but the identity of this metabolite could not be fully confirmed yet. Although the fraction also contains a methyl group at $\delta_H$ 2.81, $\delta_C$ 39.6 (presumably $CH_3$-1'') with a matching integral ratio, we were not able to connect the two spin-systems due to the low concentration of the metabolite and the resulting low S/N in selective $^1H$-NOESY experiments.

**Compound 7**

Multi-dimensional NMR spectroscopic analysis of the semipurified compound **7** ($^1H^1H$-COSY, $^1H^1H$-TOCSY allowed for the assembly of two spin systems: $\delta_H$ 6.27, CH-1'; $\delta_H$ 4.94, CH-2'; $\delta_H$ 5.36, CH-3'; $\delta_H$ 3.40, $CH_2$-5'. Selective 1D $^1H$-NOESY spectra originating from $\delta_H$ 6.27, CH-1' allowed for successful connection of the furanose moiety with the cytosine residue: $\delta_H$ 7.49, CH-6; 6.03, CH-5. Additional selective 1D $^1H$-NOESY originating from $\delta_H$ 3.40, $CH_2$-5' revealed the terminal methyl group $CH_3$-1'' ($\delta_H$ 2.15). Indicating a strong structural homology to metabolite **6**. LC-ESI-MS analysis of the sample containing enriched **7** in positive ion mode afforded the molecular ion $[M+H]^+$ of 256.0747, indicating a molecular formula of $C_{10}H_{14}N_3O_3S$ ($m/z_{calc}$ = 256.0747, $m/z_{found}$ = 256.0750), implying the detection of the reduced analogue of **6**. Evaluation of the combined spectroscopic and spectrometric data allowed for the putative assignment of **7** as the previously unreported 3',5'-dideoxy-3',4'-didehydrocytidine-5'-methylsulfinyl (ddhC-5'MeS, **7**).

**Compound 8**

Analysis of the early eluting preparative LC fractions 1 and 2 (eluting at 0-4 minutes) revealed the presence of additional ddhN species. Two dimensional homonuclear NMR spectroscopic analysis permitted the assembly of the spin system: $\delta_H$ 6.33, CH-1'; $\delta_H$ 5.08, CH-2'; $\delta_H$ 5.60, CH-3', attributed to compound **8**. LC-MS analysis of the fractions containing compound **8** monitoring for precursor ions leading to cytidine fragmentation indicated a molecular formula: $[M+H]^+$ of $C_{14}H_{21}N_4O_5S$, ($m/z_{calc}$ = 357.1226, $m/z_{found}$ = 357.1227). We note that in the case of metabolite **8**, in-source fragmentation of cytidine (*m/z* = 112.0511, calculated for $C_4H_6N_3O$) was not observed, and only seen under comparatively “higher energy” collision induced dissociation conditions, this presumably due to positive charge localisation onto the sulfonium portion of compound **8**, rather than the nucleobase moiety of compounds **1-7**, precluding facile pinacol-type fragmentation as observed in source for other metabolites within the series. Evaluation of the combined spectroscopic evidence allowed for the putative assignment of **8** as the previously unreported 3',5'-dideoxy-3',4'-didehydrocytidine-5'-Methionine (ddhC-5'Met, **8**). Direct NMR spectral comparison to an authentic standard of **8** prepared in our laboratories was thwarted by the facile decomposition of compound **8** following international transit, however UPLC-MS analysis of this mixture and comparison of retention time and MS/MS fragmentation for the presumed molecular ion allowed for the provisional validation of metabolite **8**. As with compound **6**, we note the observation of a likely epimer of compound **8**, deemed **epi-8**, deriving from hindered interconversion of the sulfonium stereocenter, with an observed spin system: $\delta_H$ 6.27, CH-1'; $\delta_H$ 5.10, CH-2'; $\delta_H$ 5.60, CH-3'. As well as an apparent peak doubling of the molecular ion attributable to **8** (and **epi-8**) in LC fractions 1 and 2. The full elucidation of **epi-6** and **epi-8** remains under investigation but is uncontroversial in light of the presented evidence.

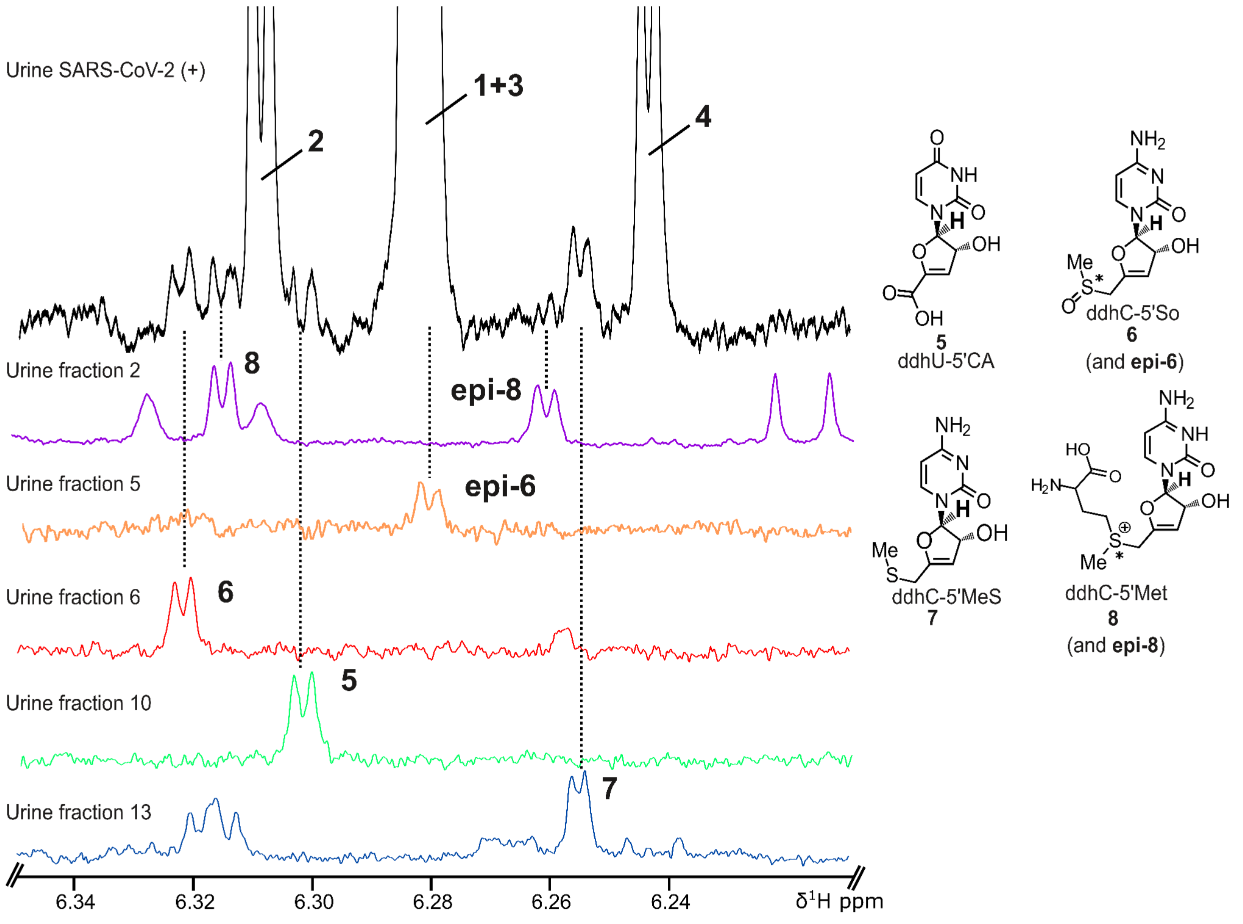


Figure S4. 1D spectra of fractions for compounds **5-8.**
Detection of 3',5'-dideoxy-3',4'-didehydrouridine-5'-carboxylic acid (ddhU-5'CA, **5**), 3',5'-dideoxy-3',4'-didehydrocytidine-5'-sulfoxide (ddhC-5'SO, **6**), 3',5'-dideoxy-3',4'-didehydrocytidine-5'-methylsulfinyl (ddhC-5'MeS, **7**) and 3',5'-dideoxy-3',4'- didehydrocytidine-5'-Methionine (ddhC-5'Met, **8**). Excerpt of the proton NMR spectrum of SARS-CoV-2 (+) subject with high concentrations of ddhN (black) with the spectra from fractions of the same urine after LC separation containing and highlighting the anomeric H-1' position of **5** (green), **6** (red), **7** (blue), and **8** (purple). Additionally, the likely epimers **epi-6** (orange) and **epi-8** (purple; same fraction as **8**) are highlighted.

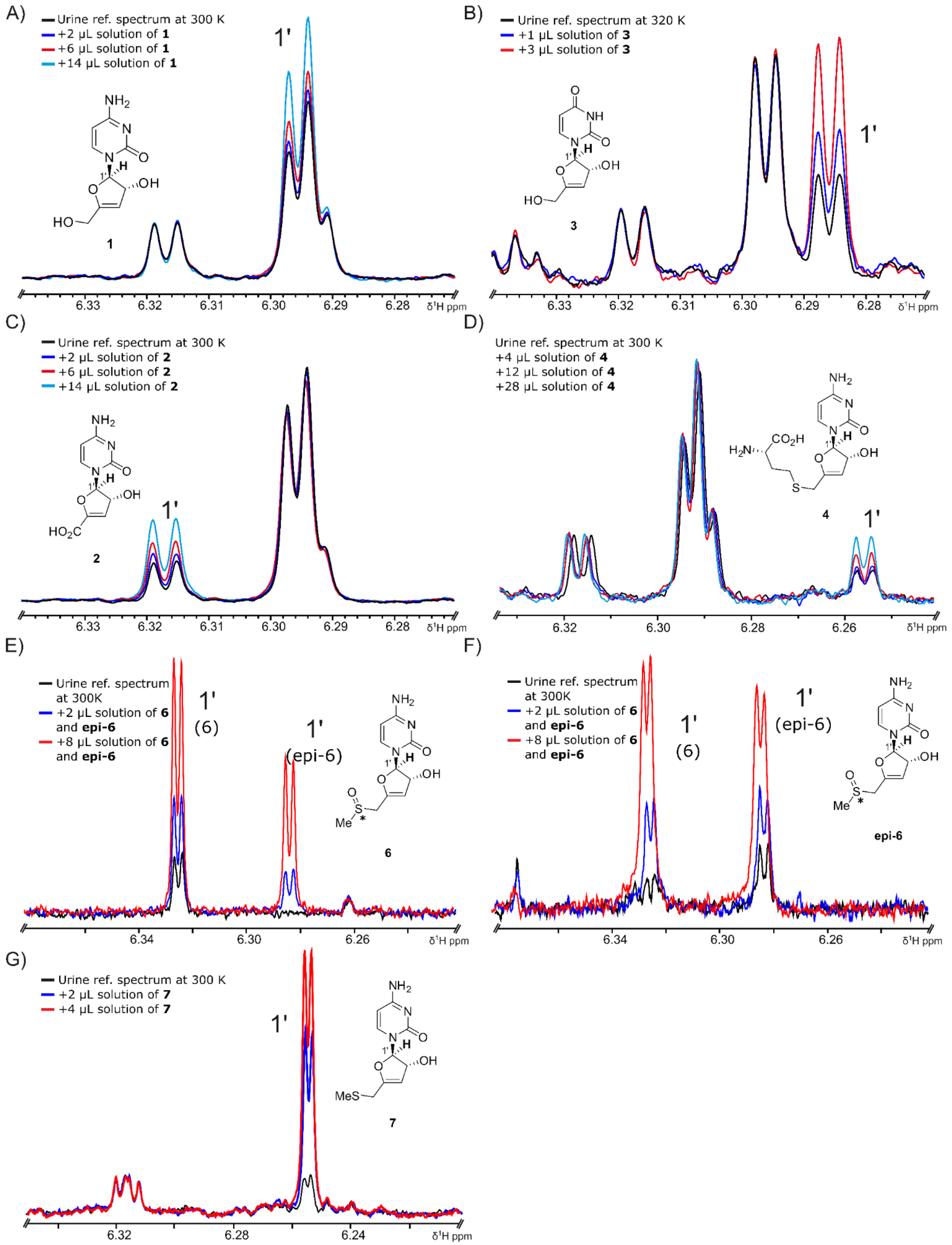

A)
Urine ref. spectrum at 300 K
+2 µL solution of 1
+6 µL solution of 1
+14 µL solution of 1
1'
1
6.33
6.32
6.31
6.30
6.29
6.28
δ¹H ppm
B)
Urine ref. spectrum at 320 K
+1 µL solution of 3
+3 µL solution of 3
1'
3
6.33
6.32
6.31
6.30
6.29
6.28
δ¹H ppm
C)
Urine ref. spectrum at 300 K
+2 µL solution of 2
+6 µL solution of 2
+14 µL solution of 2
1'
2
6.33
6.32
6.31
6.30
6.29
6.28
δ¹H ppm
D)
Urine ref. spectrum at 300 K
+4 µL solution of 4
+12 µL solution of 4
+28 µL solution of 4
1'
4
6.32
6.30
6.28
6.26
δ¹H ppm
E)
Urine ref. spectrum at 300K
+2 µL solution of 6 and epi-6
+8 µL solution of 6 and epi-6
1' (6)
1' (epi-6)
6
6.34
6.30
6.26
δ¹H ppm
F)
Urine ref. spectrum at 300K
+2 µL solution of 6 and epi-6
+8 µL solution of 6 and epi-6
1' (6)
1' (epi-6)
epi-6
6.34
6.30
6.26
δ¹H ppm
G)
Urine ref. spectrum at 300 K
+2 µL solution of 7
+4 µL solution of 7
1'
7
6.32
6.28
6.24
δ¹H ppm

Figure S5. Spiking of synthetic standards for structural confirmation.
A) Addition of metabolite **1** in increasing concentrations to a urine sample at 300K. B) Addition of metabolite **2** in increasing concentrations to a urine sample at 300K. C) Addition of metabolite **3** in increasing concentrations to a urine sample at 320K. At 320K the peaks of metabolite **1** and **3** are deconvoluted allowing the observation of an unambiguous increase of metabolite **3**. D) Addition of metabolite **4** in increasing concentrations to a urine sample at 300K. E) Addition of a metabolite solution containing **6** and **epi-6** in increasing concentration to fractionated urine (fraction 6 of figure S4) at 300 K. F) Addition of a metabolite solution containing **6** and **epi-6** in increasing concentration to fractionated urine (fraction 5 of figure S4) at 300 K. G) E) Addition of metabolite **7** in increasing concentration to fractionated urine (fraction 13 of figure S4) at 300 K.

**Spectra assignments**

Table S1A. Chemical shifts and assignment of new biomarkers **1-3**.
Urine: phosphate buffer, T = 300 K, $^{1}H$ = 800 MHz, $^{13}C$ = 200 MHz

| | **1** | | **2** | | **3** | |
|---|---|---|---|---|---|---|
| *No.* | $^{1}H$, m, *J* in Hz | $^{13}C$, type | $^{1}H$, m, *J* in Hz | $^{13}C$, type | $^{1}H$, m, *J* in Hz | $^{13}C$, type |
| *2* | | 160.1, C | | 159.7, C | | 154.6, C |
| *4* | | 169.1, C | | 169.5, C | | 169.1, C |
| *5* | 6.02, d, 7.4 | 99.4, CH | 6.02, d, 7.4 | 99.4, CH | 5.84, d, 8.3 | 105.4, CH |
| *6* | 7.37, d, 7.4 | 143.7, CH | 7.37, d, 7.4 | 144.7, CH | 7.43, d, 8.3 | 144.4, CH |
| *1'* | 6.29, d, 1.9 | 96.5, CH | 6.31, d, 2.1 | 97.1, CH | 6.29, d, 1.9 | 95.7, CH |
| *2'* | 4.93, m | 81.4, CH | 5.01, m | 82.4, CH | 5.02, m | 81.2, CH |
| *3'* | 5.38, d, 2.9 | 103.5, CH | 5.89, d, 2.8 | 110.5, CH | 5.40, m | 103.2, CH |
| *4'* | | 164.5, C | | 158.8, C | | 164.5, C |
| *5'α* | 4.29, m | 58.6, $CH_2$ | | n.d. | 4.29, m | 58.6, $CH_2$ |
| *5'β* | 4.27, m | | | | 4.27, m | |

Table S1B. Chemical shifts and assignment of new biomarkers **4** and **5**.
Reconstituted urine: phosphate buffer, T = 300 K, $^{1}H$ = 800 MHz, $^{13}C$ = 200 MHz

| | **4** | | **5** | |
|---|---|---|---|---|
| *No.* | $^{1}H$, m, J in Hz | $^{13}C$, type | $^{1}H$, m, J in Hz | $^{13}C$, type |
| *2* | | 158.4, C | | nd |
| *4* | | 167.8, C | | nd |
| *5* | 6.04, d, 7.4 | 99.4, CH | 5.87, d, 8.3 | nd |
| *6* | 7.48, d, 7.4 | 144.1, CH | 7.44, d, 8.3 | nd |
| *1'* | 6.26, d, 2.0 | 96.7, CH | 6.31, d, 2.4 | 95.9, CH |
| *2'* | 4.99, m | 81.4, CH | 5.13, dd, 2.4, 2.9 | nd |
| *3'* | 5.37, m | 104.5, CH | 5.91, d, 2.9 | nd |
| *4'* | | 161.0, C | | nd |
| *5'α* | 3.49, d, 14.8 | 29.7, $CH_2$ | | nd |
| *5'β* | 3.44, d, 14.8 | | | |
| *1''* | 3.85, m | 56.9, CH | | |
| *2''α* | 2.14, m | 32.9, $CH_2$ | | |
| *2''β* | 2.19, m | | | |
| *3''α* | 2.71, m | 29.5, $CH_2$ | | |
| *3''β* | 2.72, m | | | |

Table S1C. Chemical shifts and assignment of new biomarkers **6** and **7**.
Reconstituted urine: phosphate buffer, T = 300 K, $^{1}H$ = 800 MHz, $^{13}C$ = 200 MHz

| | **6** | | **7** | |
|---|---|---|---|---|
| *No.* | $^{1}H$, m, J in Hz | $^{13}C$, type | $^{1}H$, m, J in Hz | $^{13}C$, type |
| *2* | | nd | | nd |
| *4* | | nd | | nd |
| *5* | 6.04, d, 7.4 | nd | 6.02, d, 7.4 | nd |
| *6* | 7.47, d, 7.4 | nd | 7.48, d, 7.4 | nd |
| *1'* | 6.33, d, 2.2 | 96.7, CH | 6.27, d, 1.7 | 96.2, CH |
| *2'* | 5.05, m | nd | 4.94, m | nd |
| *3'* | 5.56, d, 2.8 | nd | 5.36, d, 2.7 | nd |
| *4'* | | nd | | nd |
| *5'α* | 4.00, m | nd | 3.28, m | nd |
| *5'β* | 3.87, m | | 3.28, m | |
| *1''* | 2.81, s | 39.6, $CH_3$ | 2.15, s | nd |

Table S2. Chemical shift of reference compounds **1** to **3**.

| | Gizzi, *Nature*, **2018**. $D_2O$, 298 K, 600 MHz, 150 MHz | | Petrova, *Tet Lett*, **2010**. $D_2O$, 300 K, 600 MHz, 150 MHz | | | |
|---|---|---|---|---|---|---|
| | **1** | | **1** | | **3** | |
| *No.* | $^1$H | $^{13}$C | $^1$H | $^{13}$C | $^1$H | $^{13}$C |
| *2* | | 159.9 | | 159.93 | | 154.23 |
| *4* | | 169.0 | | 169.00 | | 169.19 |
| *5* | 5.99 | 99.1 | 5.99 | 99.14 | 5.86 | 105.44 |
| *6* | 7.38 | 143.6 | 7.38 | 143.6 | 7.44 | 144.10 |
| *1'* | 6.27 | 96.4 | 6.27 | 96.44 | 6.29 | 95.80 |
| *2'* | 4.93 | 81.4 | 4.93 | 81.39 | 5.02 | 82.24 |
| *3'* | 5.35 | 103 | 5.35 | 102.97 | 5.40 | 103.20 |
| *4'* | | 163.9 | | 163.92 | | 164.19 |
| *5' α* | 4.29 | 58.9 | 4.29 | 58.93 | 4.30 | 59.03 |
| *5' β* | 4.26 | | 4.26 | | 4.27 | |

### Syntheses

In order to unambigiously confirm the identity of metabolites **1**-**4**, **6** and **7**, synthetic samples of each were prepared for use in spiking experiments. ddhC (**1**) was prepared according to previously reported methods[9], while ddhU (**3**) was prepared by silyl ether cleavage of known intermediate **S1** (Figure S6A)[10]. The spectroscopic data for these compounds were in good agreement with those previously reported[8]. ddhC-5'CA (**2**) was prepared from known ddhC (**1**) derivative 4-*N*-benzoyl-2′-*O*-*tert*-butyldimethylsilyl-ddhC (**S2**)[9] by sequential oxidation and global deprotection as shown in Figure S6B. ddhC-5'Hcy (**4**) was prepared according to the recently reported method[11]. Sulfide **7** was prepared from known chloride **S5**[11] by substitution with methanethiolate, then global deprotection as shown in Figure S6C. Oxidation of sulfide **S7** afforded a diastereomeric mixture of sulfoxides **S8**, along with sulfone **S9**. Deprotection of sulfoxides **S8** afforded sulfoxides **6**, which were also an inseparable diastereomeric mixture.

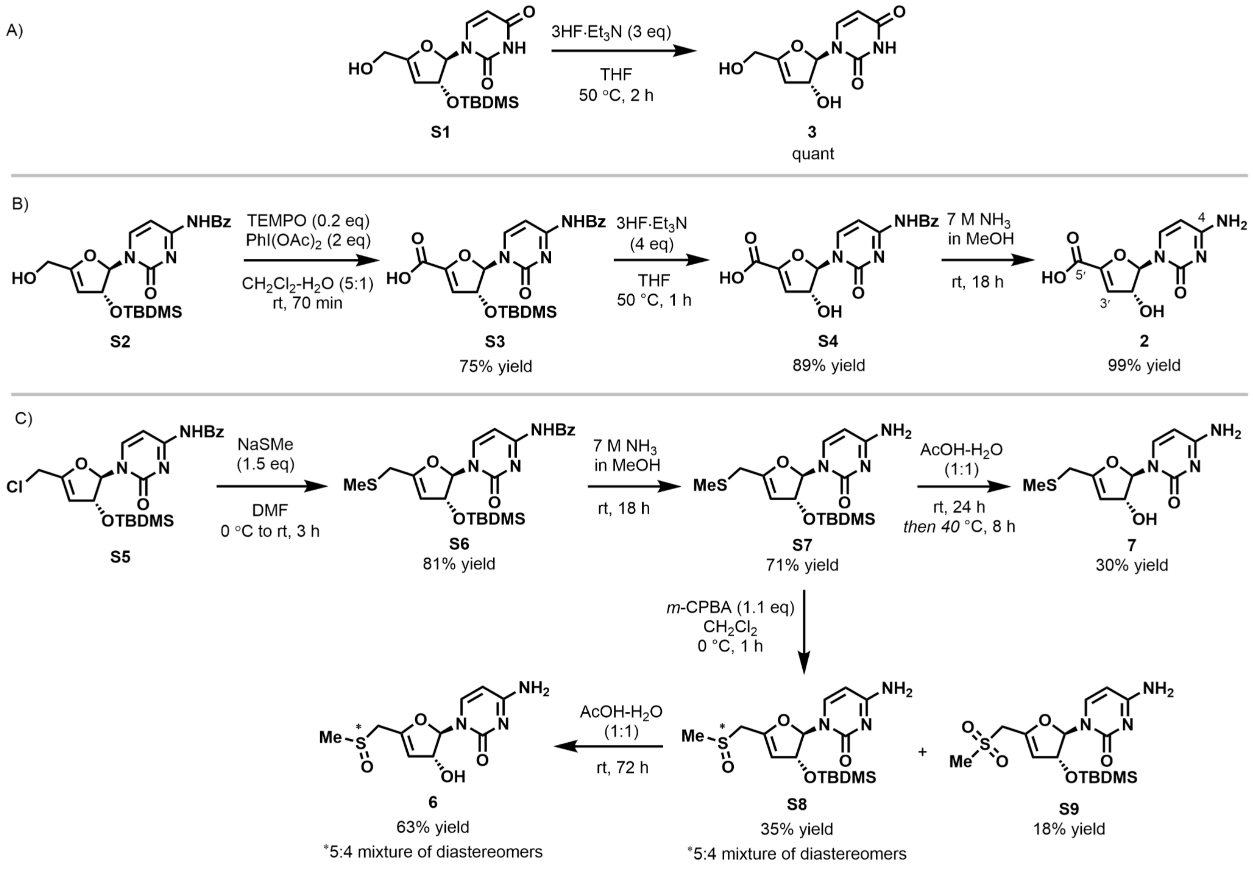


Figure S6: Synthesis of ddhN standards **2**,**3**, **6** and **7** for $^{1}$H NMR spike in experiments

The $^{13}$C NMR spectrum of **2** in $D_2O$ contains only 8 distinct signals, presumably due to overlap of the C4 and C5′ resonances at 165.9 ppm. Evidence of the α,β-unsaturated carboxylic acid fragment by $^{1}$H-$^{13}$C-HMBC analysis was also absent, as no correlation between C3′-H to C5′

was observed. We therefore obtained the $^{13}C$ NMR spectrum of **2** in DMSO-$d_6$, where an upfield shift of the C5′ resonance to 162.3 ppm provided resolution of all 9 $^{13}C$ environments.

*3'4'-Didehydro-3'-deoxyuridine* (**3**)

To a solution of silyl ether **S1** (60 mg, 0.18 mmol) in THF (1.8 mL) at room temperature was added triethylamine trihydrofluoride (50 µL, 0.30 mmol). The reaction mixture was stirred for 1 h, then triethylamine trihydrofluoride (40 µL, 0.24 mmol) was added and the reaction heated at 50 °C for 2 h. The reaction mixture was cooled to room temperature and concentrated *in vacuo*. The residue was purified by flash column chromatography on silica gel (0–30% MeOH-EtOAc) to obtain the title compound (42 mg, quant) as a colourless solid. $^{1}H$ NMR (500 MHz, $D_2O$) δ 7.46 (d, $J$ = 8.1 Hz, 1H), 6.31 (d, $J$ = 2.0 Hz, 1H), 5.89 (d, $J$ = 8.1 Hz, 1H), 5.42 (dt, $J$ = 2.7, 1.0 Hz, 1H), 5.05 (ddt, $J$ = 2.9, 2.0, 1.0 Hz, 1H), 4.36–4.27 (m, 2H); $^{13}C$ NMR (126 MHz, $D_2O$) δ 166.2, 161.3, 151.3, 141.3, 102.6, 100.4, 93.0, 78.4, 56.2.

*4-N-Benzoyl-2'-O-(tert-butyldimethylsilyl)-3'-deoxy-3',4'-didehydrocytidine-5'-carboxylic acid* (**S3**)

To a solution of alcohol **S2** (101 mg, 0.228 mmol) and diacetoxyiodobenzene (154 mg, 0.469 mmol) in $CH_2Cl_2$-$H_2O$ (5:1, 3.0 mL) at room temperature was added TEMPO (8.0 mg, 51 µmol) and the resulting mixture was stirred vigorously for 70 min. The reaction mixture was partitioned between EtOAc (20 mL) and 0.5 M aq KOH (10 mL). The organic phase was extracted with 0.5 M aq KOH (2 × 10 mL), then the combined aqueous phases were washed with EtOAc (10 mL). The aqueous layer was cooled to 0 °C and acidified to approximately pH 2 with 3 M aq HCl, upon which the product precipitated as a colourless solid. The acidified aqueous layer was extracted with EtOAc (3 × 20 mL), then the combined organic extracts were washed with brine (10 mL), dried over anhydrous $MgSO_4$, filtered and concentrated *in vacuo* to afford the title compound (78 mg, 75% yield) as a colourless solid. $^{1}H$ NMR (500 MHz, DMSO-$d_6$) δ 8.04–7.98 (m, 2H), 7.95 (d, $J$ = 7.5 Hz, 1H), 7.67–7.60 (m, 1H), 7.55–7.47 (m, 2H), 7.35 (d, $J$ = 7.5 Hz, 1H), 6.24 (d, $J$ = 2.9 Hz, 1H), 6.03 (d, $J$ = 2.7 Hz, 1H), 5.26 (t, $J$ = 2.8 Hz, 1H), 0.86 (s, 9H), 0.10 (s, 3H), 0.08 (s, 3H); $^{13}C$ NMR (126 MHz, DMSO) δ 167.6, 163.6, 160.3, 153.8, 150.6, 145.7, 133.1, 132.8, 128.5, 128.4, 111.9, 97.1, 95.4, 79.3, 25.6, 17.7, –4.8, –4.8; HRMS (ESI/QTOF) *m/z*: $[M - H]^-$ Calcd for $C_{22}H_{26}N_3O_6Si$ 456.1591; Found: 456.1603.

*4-N-Benzoyl-3'-deoxy-3',4'-didehydrocytidine-5'-carboxylic acid* (**S4**)

To a solution of silyl ether **S3** (57 mg, 0.13 mmol) in THF (1.3 mL) at room temperature was added 3HF·$Et_3N$ (40 µL, 0.24 mmol). The reaction mixture was stirred at room temperature for 1 h, then treated with additional 3HF·$Et_3N$ (50 µL, 0.30 mmol) and heated to 50 °C for 1 h. The reaction mixture was then cooled to room temperature and concentrated *in vacuo*. Purification by flash column chromatography on C18 silica gel (5–100% MeOH-$H_2O$) afforded the title compound (38 mg, 89% yield) as a colourless solid after lyophilization from water. $^1$H NMR (500 MHz, DMSO-$d_6$) δ 11.27 (s, 1H), 8.03–7.98 (m, 2H), 7.73 (d, *J* = 7.5 Hz, 1H), 7.65–7.59 (m, 1H), 7.55–7.48 (m, 2H), 7.33 (d, *J* = 7.3 Hz, 1H), 6.16 (d, *J* = 2.1 Hz, 1H), 5.67 (d, *J* = 6.5 Hz, 1H), 5.44 (br s, 1H), 4.71 (d, *J* = 4.4 Hz, 1H); $^{13}$C NMR (126 MHz, DMSO) δ 167.5, 163.2, 162.1, 157.5, 154.1, 144.5, 133.1, 132.8, 128.5, 128.5, 105.6, 96.7, 93.9, 78.5; HRMS (ESI/QTOF) *m/z*: $[M + Na]^+$ Calcd for $C_{16}H_{13}N_3O_6Na$ 366.0702; Found: 366.0692.

*3'-Deoxy-3',4'-didehydrocytidine-5'-carboxylic acid* (**2**)

Benzoyl amide **S4** (43 mg, 0.13 mmol) was dissolved in methanolic ammonia (7 M, 1 mL) and stirred at room temperature overnight. The reaction mixture was concentrated *in vacuo*, then the residue purified by passage through a plug of silica gel, eluting with 0–50% $H_2O$-MeCN containing 1% conc $NH_4OH$. Lyophilization from water afforded the title compound (30 mg, 99% yield) as a colourless solid. $^1$H NMR (500 MHz, DMSO-$d_6$) δ 7.32 (br s, 1H), 7.20 (d, J = 7.5 Hz, 1H), 6.16 (d, J = 2.7 Hz, 1H), 5.74 (d, J = 7.4 Hz, 1H), 5.53 (d, J = 2.6 Hz, 1H), 4.71 (t, J = 2.7 Hz, 1H); $^{13}$C NMR (126 MHz, DMSO) δ 166.1, 162.3, 156.6, 155.2, 141.2, 107.2, 95.3, 93.8, 78.6; $^1$H NMR (500 MHz, $D_2O$) δ 7.41 (d, J = 7.5 Hz, 1H), 6.33 (d, J = 2.3 Hz, 1H), 6.04 (d, J = 7.5 Hz, 1H), 5.92 (d, J = 2.8 Hz, 1H), 5.03 (dd, J = 2.8, 2.2 Hz, 1H); $^{13}$C NMR (126 MHz, $D_2O$) δ 166.0, 165.9, 156.7, 155.1, 141.1, 107.5, 96.4, 93.8, 79.0; HRMS (ESI/QTOF) m/z: $[M - H]^-$ Calcd for $C_9H_8N_3O_5$ 238.0464; Found: 238.0469.

*4-N-Benzoyl-2'-O-(tert-butyldimethylsilyl)-3',4'-didehydro-3',5'-dideoxy-5'-(methyl)thio-cytidine* (**S6**)

A stream of argon was bubbled through a solution of chloride **S5** (144 mg, 0.312 mmol) in anhydrous DMF (3 mL) for 30 min. The degassed solution was cooled to 0 °C under an atmosphere of argon and sodium methanethiolate (37 mg, 0.50 mmol) was added in one portion. The solution was stirred at 0 °C for 15 min, before being allowed to warm to room temperature. After 3 h, the reaction was quenched with AcOH (28 µL, 0.47 mmol), then concentrated *in vacuo* at room temperature. The resulting residue was purified by flash column chromatography on silica gel (10–50% (3:1 *v/v* EtOAc-EtOH)-petroleum ether) to afford sulfide **S6** (120 mg, 81% yield) as a light yellow solid. $^1$H NMR (500 MHz, $CDCl_3$) δ 8.69 (br s, 1H), 7.93–7.85 (m, 3H), 7.64–7.45 (m, 4H), 6.30 (d, *J* = 1.3 Hz, 1H), 5.08 (dt, *J* = 2.5, 0.8 Hz, 1H), 4.83–4.81 (m, 1H), 3.31 (s, 1H), 2.17 (s, 3H), 0.91 (s, 9H), 0.18 (s, 3H), 0.11 (s, 3H); $^{13}$C NMR (126 MHz, $CDCl_3$) δ 162.5, 158.2, 143.9, 133.4, 129.8, 129.3, 127.6, 102.5, 94.6, 81.0, 30.8, 25.9, 18.3, 16.1, −4.4, −4.7. HRMS (ESI/QTOF) *m/z*: $[M + Na]^+$ Calcd for $C_{23}H_{31}N_3O_4NaSiS$ 496.1702; Found: 496.1700.

*2'-O-(tert-Butyldimethylsilyl)-3',4'-didehydro-3',5'-dideoxy-5'-(methyl)thio-cytidine* (**S7**)

Benzoyl amide **S6** (110 mg, 0.232 mmol) was dissolved in methanolic ammonia (7 M, 3 mL) and the reaction mixture was stirred at room temperature overnight. The reaction mixture was concentrated *in vacuo*, and the residue obtained was purified by flash column chromatography on silica gel (20–80% (3:1 *v/v* EtOAc-EtOH)-petroleum ether) to afford the title compound **S7** (61 mg, 71% yield) as a colourless foam. $^1$H NMR (500 MHz, $CDCl_3$) δ 7.43 (d, *J* = 7.4 Hz, 1H), 6.29 (d, *J* = 1.6 Hz, 1H), 5.05 (dt, *J* = 2.5, 0.8 Hz, 1H), 4.82 (ddt, *J* = 2.5, 1.5, 0.7 Hz, 1H), 3.31–3.23 (m, 2H), 2.16 (s, 3H), 0.89 (s, 9H), 0.15 (s, 3H), 0.09 (s, 3H) ($NH_2$ protons not observed); $^{13}$C NMR (126 MHz, $CDCl_3$) δ 165.5, 158.4, 155.3, 140.9, 102.3, 94.5, 93.9, 80.9, 30.8, 25.9, 18.3, 16.1, −4.3, −4.7; HRMS (ESI/QTOF) *m/z*: $[M + H]^+$ Calcd for $C_{16}H_{28}N_3O_3SiS$ 370.1621; Found: 370.1625.

*3',4'-Didehydro-3',5'-dideoxy-5'-(methyl)thio-cytidine* (**7**)

Silyl ether **S7** (30 mg, 81 µmol) was dissolved in acetic acid-water (1:1 *v/v*, 2 mL) and the solution was stirred at room temperature overnight. The mixture was then heated to 40 °C and stirring continued for an additional 8 h. The solution was diluted with toluene (10 mL) and concentrated *in vacuo*, after which purification by flash column chromatography on silica gel (10–50% MeOH-$CH_2Cl_2$) afforded the title compound **7** (6 mg, 29% yield) as a colourless solid after a final lyophilisation from water. $^1$H NMR (500 MHz, MeOD) δ 7.50 (d, *J* = 7.5 Hz, 1H), 6.26 (d, *J* = 1.9 Hz, 1H), 5.88 (d, *J* = 7.5 Hz, 1H), 5.19 (dt, *J* = 2.6, 0.8 Hz, 1H), 4.77–4.75 (m, 1H), 3.35 (d, *J* = 14.9 Hz, 1H), 3.29 (d, *J* = 14.9 Hz, 1H, overlapping with MeOD), 2.16 (s, 3H); $^{13}$C NMR (126 MHz, MeOD) δ 167.7, 160.5, 157.9, 141.4, 102.2, 96.4, 95.3, 80.7, 31.0, 15.9; HRMS (ESI/QTOF) *m/z*: $[M + H]^+$ Calcd for $C_{10}H_{14}N_3O_3S$ 256.0756; Found: 256.0751.

*2'-O-(tert-Butyldimethylsilyl)-3',4'-didehydro-3',5'-dideoxy-5'-(methyl)sulfinyl-cytidine* (**S8**) and *2'-O-(tert-Butyldimethylsilyl)-3',4'-didehydro-3',5'-dideoxy-5'-(methyl)sulfonyl-cytidine* (**S9**)

*m*-CPBA (41 mg, 70–86% purity, 0.17–0.20 mmol) was added in one portion to a stirred solution of sulfide **S7** (63 mg, 90% purity, 0.15 mmol) in $CH_2Cl_2$, which was maintained at 0 °C under an atmosphere of argon. The reaction mixture was stirred for 1 h, then warmed to room temperature and concentrated *in vacuo*. Purification by flash column chromatography on silica gel (0–50% MeOH-$CH_2Cl_2$) afforded two products, **S8** (21 mg, 36% yield), and **S9** (18 mg, 29% yield). Sulfoxide **S8** was isolated as a 5:4 inseparable mixture of diastereomers. *Major diastereomer*: $^1$H NMR (500 MHz, $CDCl_3$) δ 7.26 (d, *J* = 7.4 Hz, 1H), 6.31 (d, *J* = 2.3 Hz, 1H), 5.84 (d, *J* = 7.4 Hz, 1H), 5.31 (d, *J* = 2.5 Hz, 1H), 4.97 (apparent t, *J* = 2.4 Hz, 1H), 3.64 (d, *J* = 13.3 Hz, 1H), 3.57 (d, *J* = 13.3 Hz, 1H), 2.67 (s, 3H), 0.87 (s, 9H), 0.10 (s, 3H), 0.07 (s, 3H); $^{13}$C NMR (126 MHz, $CDCl_3$) δ 166.0, 155.3, 151.9, 141.2, 106.8, 95.9, 95.3, 80.6, 53.5, 39.2, 25.9, 18.2, −4.4, −4.6. *Minor diastereomer*: $^1$H NMR (500 MHz, $CDCl_3$) δ 7.38 (d, *J* = 7.4 Hz, 1H), 6.17 (d, *J* = 2.7 Hz, 1H), 5.87 (d, *J* = 7.4 Hz, 1H), 5.28 (d, *J* = 2.5 Hz, 1H), 5.07 (apparent t, *J* = 2.6 Hz, 1H), 3.70 (d, *J* = 13.8 Hz, 1H), 3.54 (d, *J* = 13.8 Hz, 1H), 2.69 (s, 3H), 0.87 (s, 9H), 0.08 (s, 3H), 0.06 (s, 3H); $^{13}$C NMR (126 MHz, $CDCl_3$) δ 166.1, 155.3, 151.1, 142.2, 107.2, 96.6, 95.9,

80.2, 51.4, 38.6, 25.9, 18.2, −4.4, −4.5. HRMS (ESI/QTOF) *m/z*: $[M + Na]^+$ Calcd for $C_{16}H_{27}N_3O_4NaSiS$ 408.1389; Found: 408.1393.

Sulfone **S9**: $^1$H NMR (500 MHz, $CDCl_3$) δ 7.28 (d, *J* = 7.4 Hz, 1H), 6.26 (d, *J* = 2.6 Hz, 1H), 5.90 (d, *J* = 7.4 Hz, 1H), 5.38 (d, *J* = 2.6 Hz, 1H), 5.04 (apparent t, *J* = 2.6 Hz, 1H), 3.97 (d, *J* = 14.7 Hz, 1H), 3.92 (d, *J* = 14.7 Hz, 1H), 3.01 (s, 3H), 0.87 (s, 9H), 0.09 (s, 3H), 0.07 (s, 3H); $^{13}$C NMR (126 MHz, $CDCl_3$) δ 166.0, 155.2, 150.0, 141.6, 107.7, 96.3, 96.0, 80.2, 54.5, 42.0, 25.9, 18.2, −4.5, −4.6; HRMS (ESI/QTOF) *m/z*: $[M + H]^+$ Calcd for $C_{16}H_{28}N_3O_5SSi$ 402.1519; Found: 402.1516.

*3',4'-Didehydro-3',5'-dideoxy-5'-(methyl)sulfinyl-cytidine* (**6**)

Silyl ether **S8** (18 mg, 47 µmol) (5:4 mixture of diastereomers at sulfur) was dissolved in acetic acid-water (1:1 *v/v*, 2 mL) and the reaction mixture was stirred at room temperature for 72 h. The solution was concentrated *in vacuo*, followed by dilution with water and lyophilisation. Purification by flash column chromatography on silica gel (0–50% $H_2O$-MeCN) afforded the title compounds **6** and epi-**6** (8 mg, 61% yield) as a colourless solid after lyophilization from water. Sulfoxide **6** was isolated as a 5:4 inseparable mixture of diastereomers, assigned A (major) and B (minor). $^1$H NMR (500 MHz, MeOD) δ 7.55 (d, *J* = 7.5 Hz, 1H, *B*), 7.48 (d, *J* = 7.5 Hz, 1H, *A*), 6.37 (d, *J* = 2.3 Hz, 1H, *A*), 6.30 (d, *J* = 2.5 Hz, 1H, *B*), 5.89 (apparent d, *J* = 7.5 Hz, 1H, *A* + *B*), 5.45–5.40 (m, 1H, *A* + *B*), 3.91 (d, *J* = 13.7 Hz, 1H, *B*), 3.86 (d, *J* = 13.7 Hz, 1H, *A*), 3.79 (d, *J* = 13.7 Hz, 1H, *A*), 3.75 (d, *J* = 13.7 Hz, 1H, *B*), 2.75 (s, 3H, *B*), 2.74 (s, 3H, *A*). $^{13}$C NMR (126 MHz, MeOD) δ 167.79, 167.75, 157.84, 157.82, 153.66, 153.65, 142.7, 142.1, 107.6, 107.4, 96.9, 96.8, 96.4, 95.7, 80.5, 80.3, 52.7, 52.2, 38.7, 38.5; HRMS (ESI/QTOF) *m/z*: $[M + H]^+$ Calcd for $C_{10}H_{14}N_3O_4S$ 272.0705; Found: 272.0699.

Table S3. Detection of compounds **1**-**3** in acute phase SARS-CoV-2 by NMR spectroscopy

Detection and quantification of compounds 1-3 in acute phase SARS-CoV-2 infected urine. Detected refers to inspection of the spectra by a human and visual confirmation of compound presence. Quantification refers to successful quantification by the fitting algorithm.

| | All Samples (100%) | | |
|---|---|---|---|
| | N | Detected | Quantification |
| ddhC (**1**) | 273 (100%) | 239 (88%) | 151 (55%) |
| ddhC-5'CA (**2**) | 273 (100%) | 185 (68%) | 92 (34%) |
| ddhU (**3**) | 273 (100%) | 199 (73%) | 151 (55%) |
| | Hospitalized (21%) | | |
| | N | Detected | Quantification |
| **1** | 57 (100%) | 56 (98%) | 40 (70%) |
| **2** | 57 (100%) | 46 (81%) | 36 (63%) |
| **3** | 57 (100%) | 51 (89%) | 40 (70%) |
| | Non-Hospitalized (68%) | | |
| | N | Detected | Quantification |
| **1** | 186 (100%) | 153 (82%) | 91 (49%) |
| **2** | 186 (100%) | 117 (63%) | 48 (26%) |
| **3** | 186 (100%) | 123 (66%) | 91 (49%) |
| | NA (11%) | | |
| | N | Detected | Quantification |
| **1** | 30 (100%) | 29 (97%) | 20 (67%) |
| **2** | 30 (100%) | 24 (80%) | 8 (27%) |
| **3** | 30 (100%) | 25 (83%) | 20 (67%) |

Table S4. Detection of compounds **1**-**3** in acute phase SARS-CoV-2 by LC-MS
Detection and quantification of compounds 1-3 in acute phase SARS-CoV-2 infected urine.

| | All Samples (100%) | | |
|---|---|---|---|
| | N | Detected | Non-Detected |
| ddhC (**1**) | 273 (100%) | 255 (93%) | 18 (7%) |
| ddhC-5'CA (**2**) | 273 (100%) | 164 (60%) | 109 (40%) |
| ddhU (**3**) | 273 (100%) | 66 (24%) | 207 (76%) |
| | Hospitalized (21%) | | |
| | N | Detected | Non-Detected |
| **1** | 57 (100%) | 57 (100%) | 0 (0%) |
| **2** | 57 (100%) | 47 (82%) | 10 (18%) |
| **3** | 57 (100%) | 25 (44%) | 32 (56%) |
| | Non-Hospitalized (68%) | | |
| | N | Detected | Non-Detected |
| **1** | 186 (100%) | 170 (91%) | 16 (9%) |
| **2** | 186 (100%) | 95 (51%) | 91 (49%) |
| **3** | 186 (100%) | 33 (18%) | 153 (82%) |
| | NA (11%) | | |
| | N | Detected | Non-Detected |
| **1** | 30 (100%) | 28 (93%) | 2 (7%) |
| **2** | 30 (100%) | 22 (73%) | 8 (27%) |
| **3** | 30 (100%) | 8 (27%) | 22 (73%) |

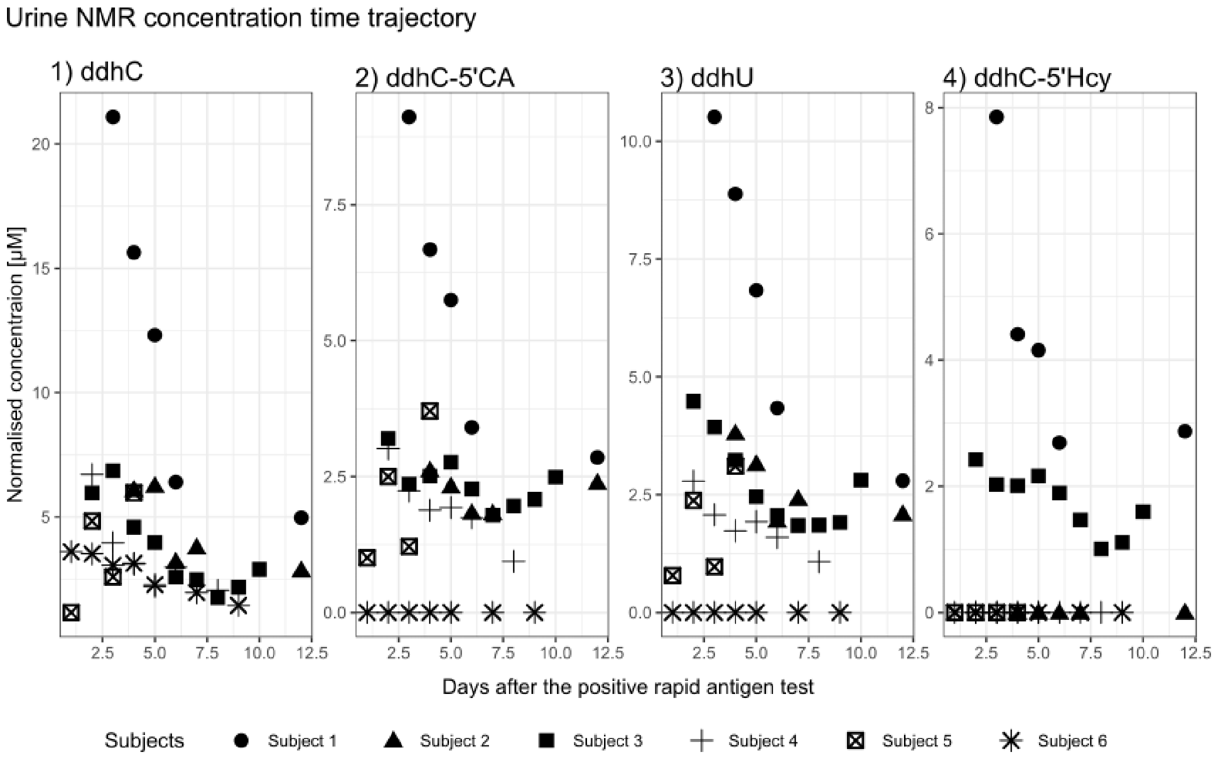


Figure S8. Longitudinal concentration data of compounds **1**-**4** from the acute SARS-CoV-2 (+) Human urine samples measured by NMR spectroscopy.

The concentration of compounds **1**-**4** is given in μM per mM creatinine. Some compounds could be detected by visual inspection but could not be quantified by line shape fitting due to low S/N e.g. **2** and **3** at time point one for subject 6.

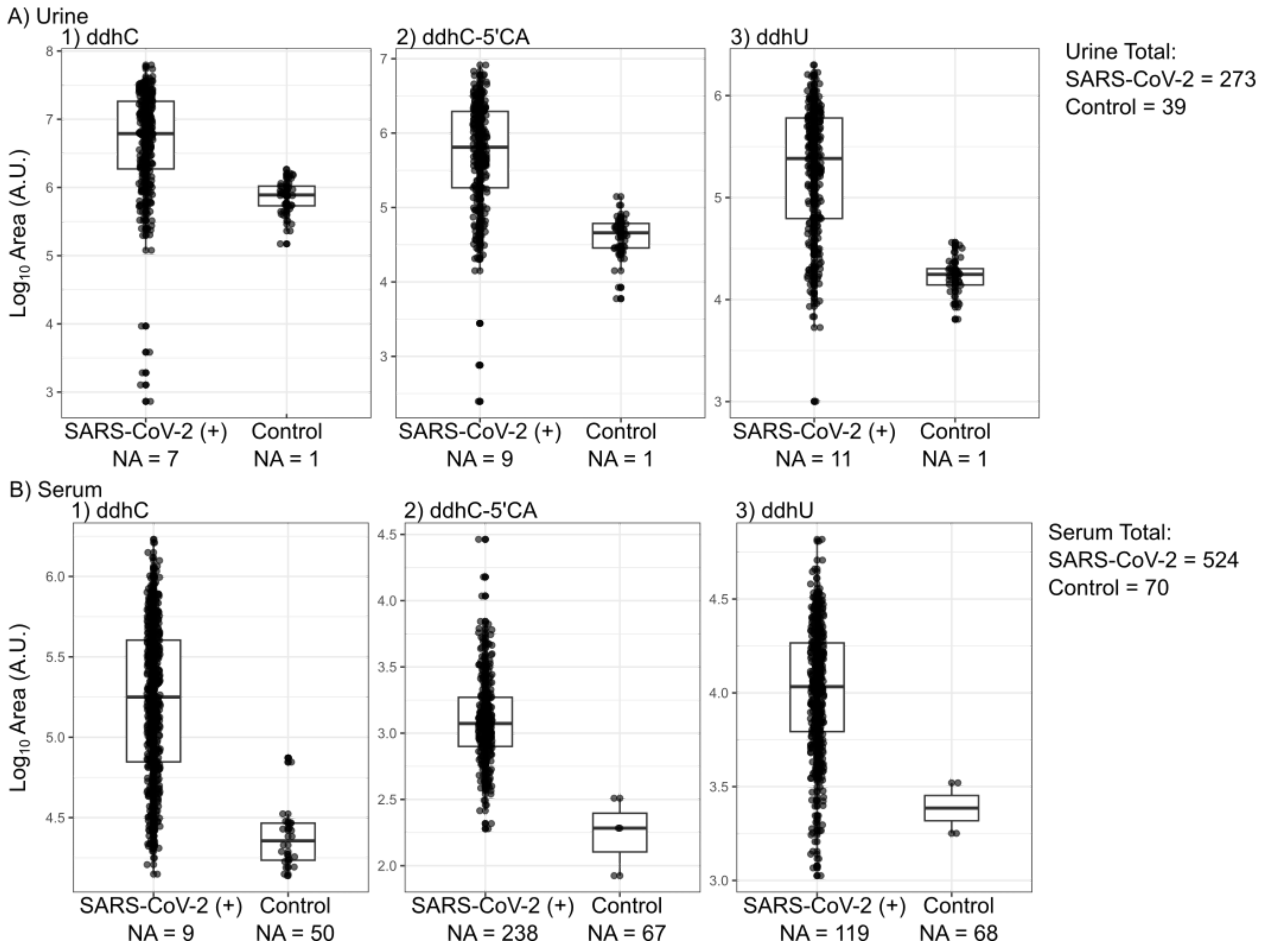


Figure S9. LC-MS peak area comparison for **1**-**4** determined in A) urine and B) serum.

Table S5 LC-QQQ-MS peak area comparison in urine and serum

| | Urine | | Serum | |
|---|---|---|---|---|
| | **SARS-CoV-2 (+)** N = 273[1] | **Control** N = 39[1] | **SARS-CoV-2 (+)** N = 524[1] | **Control** N = 70[1] |
| Area of ddhC | 10,852,730 (11,716,064) | 834,745 (422,829) | 276,485 (269,046) | 27,077 (16,556) |
| NA | 7 | 1 | 9 | 50 |
| Area of ddhC-5'CA | 1,196,885 (1,352,162) | 47,571 (26,719) | 1,712 (2,231) | 200 (120) |
| NA | 9 | 1 | 238 | 67 |
| Area of ddhU | 369,654 (371,826) | 18,284 (7,330) | 13,427 (9,651) | 2,548 (1,082) |
| NA | 11 | 1 | 119 | 68 |

[1] Mean (SD)

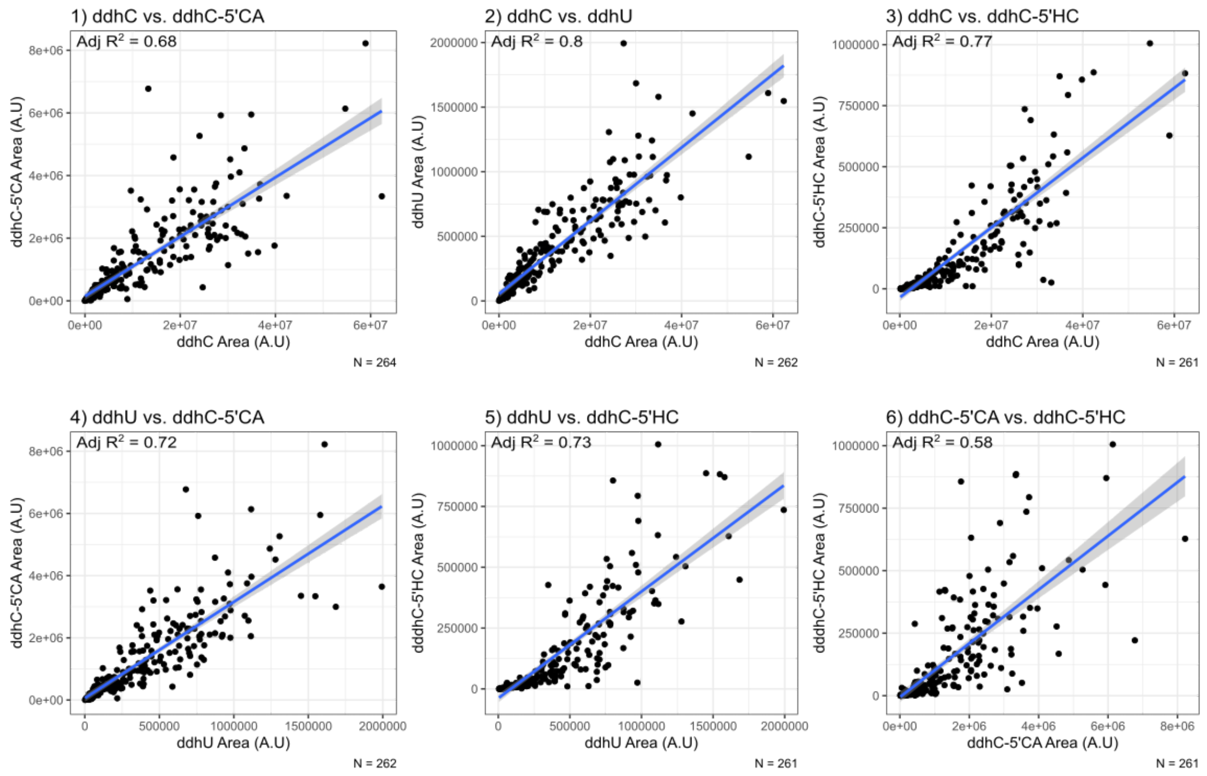


Figure S10. Area correlation for compounds **1**-**4** in urine determined by LC-QQQ-MS.

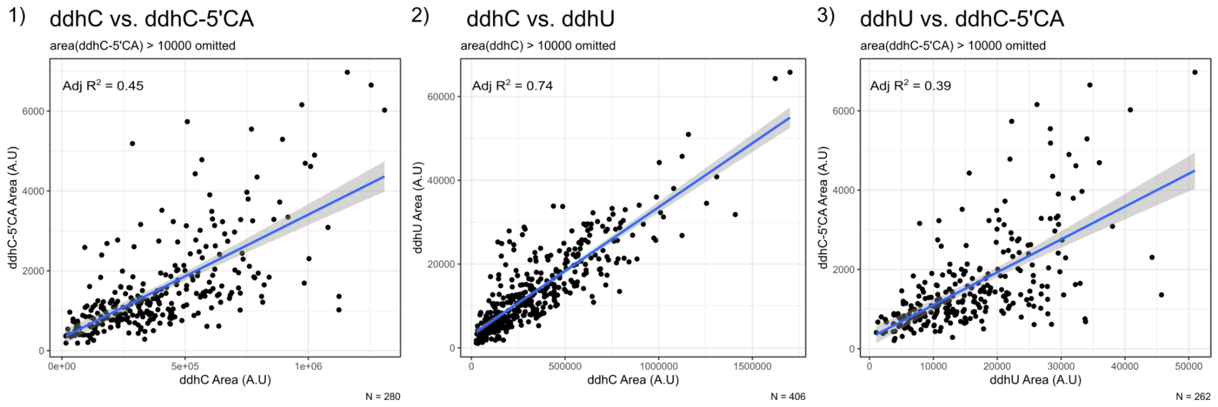


Figure S11. Area correlation for compounds **1**-**3** in serum determined by LC-QQQ-MS.

**Positive mode LC-QToF-MS/MS spectra of authentic standards**

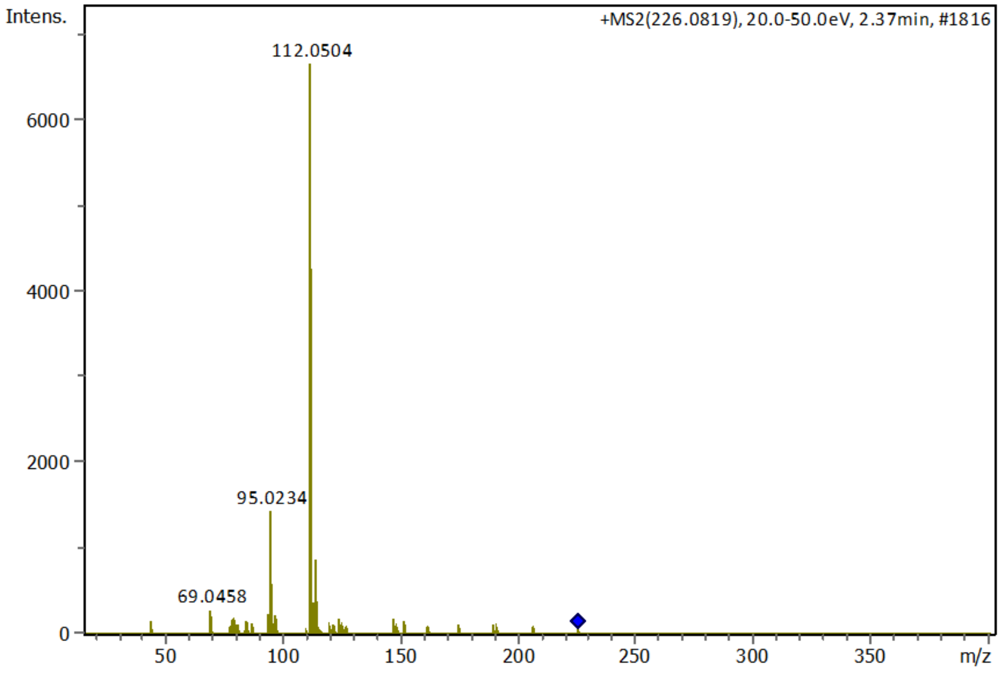


Figure S12. MS/MS spectrum for ddhC (**1**).

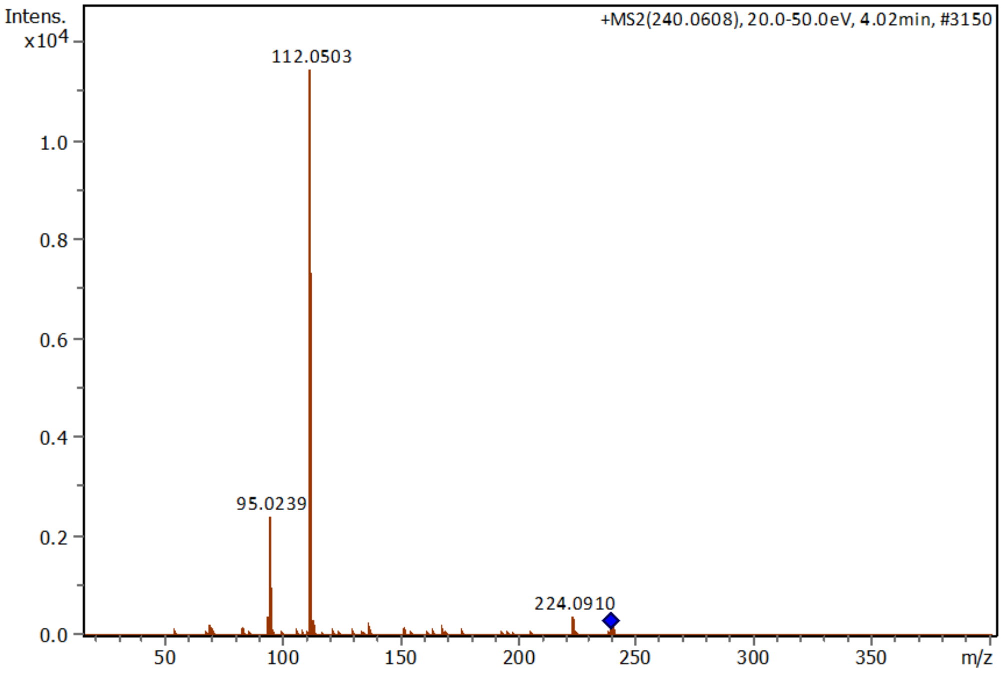


Figure S13. MS/MS spectrum for ddhC-5'CA (**2**)

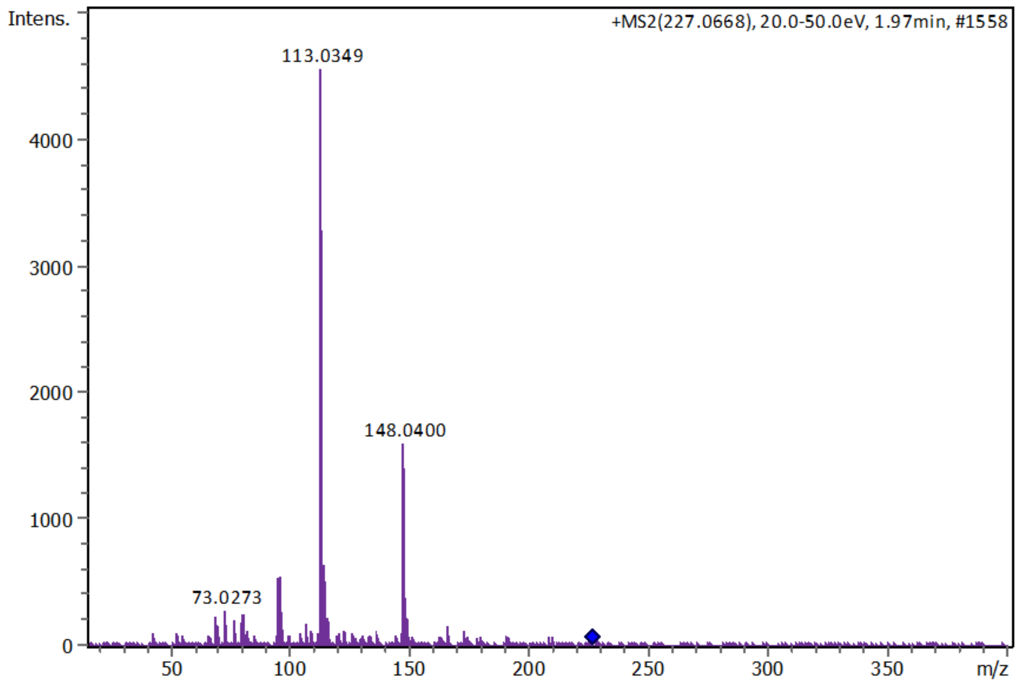


Figure S14. MS/MS spectrum for ddhU (**3**)

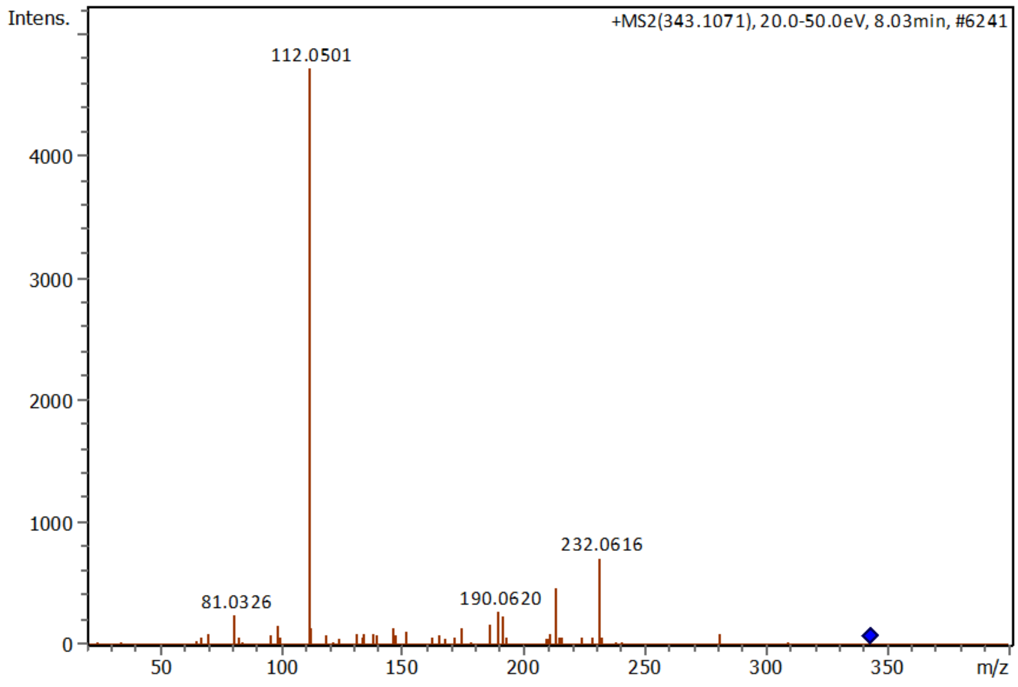


Figure S15. MS/MS spectrum for ddhC-5'Hcy (**4**)
